# Dysregulated inflammation in solid tumor malignancy patients shapes polyfunctional antibody responses to COVID-19 vaccination

**DOI:** 10.1101/2025.05.09.25327344

**Authors:** Ruth A. Purcell, Marios Koutsakos, Lukasz Kedzierski, Lilith F. Allen, Oscar H. Lloyd Williams, Jo-Wai D. Wang, George Cavic, Adam K. Wheatley, Wen Shi Lee, Bruce D. Wines, P. Mark Hogarth, Emily M. Eriksson, Ivo Mueller, Katherine A. Bond, Deborah A. Williamson, Janine M. Trevillyan, Jason A. Trubiano, Thi H.O. Nguyen, Pradhipa Ramanathan, Stephen J. Rogerson, Kelly B. Arnold, Kanta Subbarao, Adrian Lee, Amanda L. Hudson, Alexander Yuile, Helen R. Wheeler, Stephen J. Kent, Kevin John Selva, Siddhartha Mahanty, Katherine Kedzierska, Aude M. Fahrer, Yada Kanjanapan, Amy W. Chung

## Abstract

Solid tumor malignancy (STM) patients experience increased risk of breakthrough SARS-CoV-2 infection owing to reduced COVID-19 vaccine immunogenicity. However, the underlying immunological causes of impaired neutralization remain poorly characterized. Furthermore, non-neutralizing antibody functions can contribute to reduced disease severity but remain understudied within high-risk populations. We dissected polyfunctional antibody responses in STM patients and age-matched controls who received adenoviral vector- or mRNA-based COVID-19 vaccine regimens. Elevated inflammatory biomarkers, including agalactosylated IgG, interleukin (IL)-6, IL-18, and an expanded population of CD11c^−^CD21^−^ double negative 3 (DN3) B cells were observed in STM patients. These inflammatory biomarkers were associated with impaired neutralization and reduced IgG targeting the antigenically novel receptor binding domain. In contrast, mRNA vaccination induced Fc effector functions that were comparable in patients and controls and were cross-reactive against SARS-CoV-2 variants, supported by robust generation of IgG against the antigenically conserved spike 2 domain in STM patients. These data highlight the resilience of Fc functional antibodies and identify systemic inflammatory biomarkers that may underpin impaired neutralizing antibody responses, suggesting potential avenues for immunomodulation via rational vaccine design.

## INTRODUCTION

The immunogenicity of COVID-19 vaccines, as with other respiratory virus vaccines, is highly variable and typically impaired in individuals who are immunocompromised ^1, 2, 3, 4, 5^ and consequently vulnerable to severe disease outcomes^6, 7, 8^. Accordingly, there is an urgent need to optimize vaccine efficacy in high-risk populations^9, 10^, however, mechanisms underpinning impaired vaccine immunogenicity in these groups remain understudied.

Solid tumor malignancy (STM) patients are a heterogenous high-risk group^7, 8^ in whom characterization of COVID-19 vaccine responses remains largely limited to reports of impaired neutralizing and binding antibody titers^1, 2^. Neutralizing antibodies remain the best described correlate of protection against COVID-19^11, 12, 13, 14^. However, when neutralizing titers are low, for example against SARS-CoV-2 variants^15, 16, 17, 18, 19^ or in immunocompromised vaccinees^1, 2, 3, 4, 5^, maintenance of robust T cell and extra-neutralizing antibody functions may be important in mitigating disease severity and blocking transmission^14, 19, 20^. Unlike neutralizing antibodies which must target specific epitopes^21^, Fc functional antibodies may target any epitope, although induction of specific Fc functions, such as phagocytosis and cytotoxicity, as long as they are conformationally compatible with Fc gamma receptor (FcγR) engagement^22^. This ensures that despite loss of SARS-CoV-2 variant-specific antibodies against neutralizing epitopes, particularly those targeting the antigenically variable receptor binding domain (RBD)^23, 24, 25^, preservation of antibody responses against antigenically conserved Spike 2 (S2) and other non-neutralizing epitopes may facilitate robust antiviral Fc effector functions^18, 26, 27^. Consequently, Fc effector functions against SARS-CoV-2 are more durable and cross-reactive than neutralization^18, 27, 28,29^, facilitating control of infection and enhanced protection under circumstances of impaired neutralization^20, 29, 30^.

Here, we recruited infection-naive STM patients and age-matched controls who received mRNA or adenoviral vector-based COVID-19 vaccine regimens early in the pandemic. We performed in-depth characterization of vaccine-induced functional antibody responses and further probed how systemic inflammatory dysregulation experienced by STM patients^31, 32^ may modulate COVID-19 vaccine responses. We demonstrate that mRNA-based COVID-19 vaccination induces robust cross-reactive Fc functions in STM patients despite inflammation-associated impairment of neutralization and identify inflammatory biomarkers that may contribute to impaired COVID-19 vaccine immunogenicity.

## RESULTS

### Three mRNA vaccine doses overcome impaired SARS-CoV-2 neutralization breadth-potency in STM patients

Two cross-sectional cohorts of COVID-19-naive STM patients and age-matched controls who received COVID-19 vaccines encoding the ancestral SARS-CoV-2 spike were recruited. Vaccinees in the mRNA-primed cohort received two mRNA-based BNT162b2 doses (*n* = 30 control; *n* = 40 STM) plus an mRNA-based booster (*n* = 20 control; *n* = 42 STM). Vaccinees in the adenoviral vector-primed cohort received two adenoviral vector-based AZD1222 doses (*n* = 29 control; *n* = 47 STM) plus an mRNA-based booster (*n* = 21 control; *n* = 59 STM) (Fig. 1A and Supplementary Table 1). The cohorts comprised a diversity of malignancies, with breast, brain, gastrointestinal, genitourinary, and gynaecological malignancies being most frequent. Across cohorts and timepoints, between 40%-57% of vaccinees had metastatic disease. At least half of patients in the mRNA-primed cohort and at least one third of patients in the adenoviral vector-primed cohort were under an active chemotherapy regimen at the time of vaccination. Patients not on chemotherapy were receiving an immunotherapy, targeted therapy, or radiotherapy regimen and a minority were under observation following prior treatment (Supplementary Table 1).

**Fig. 1.**
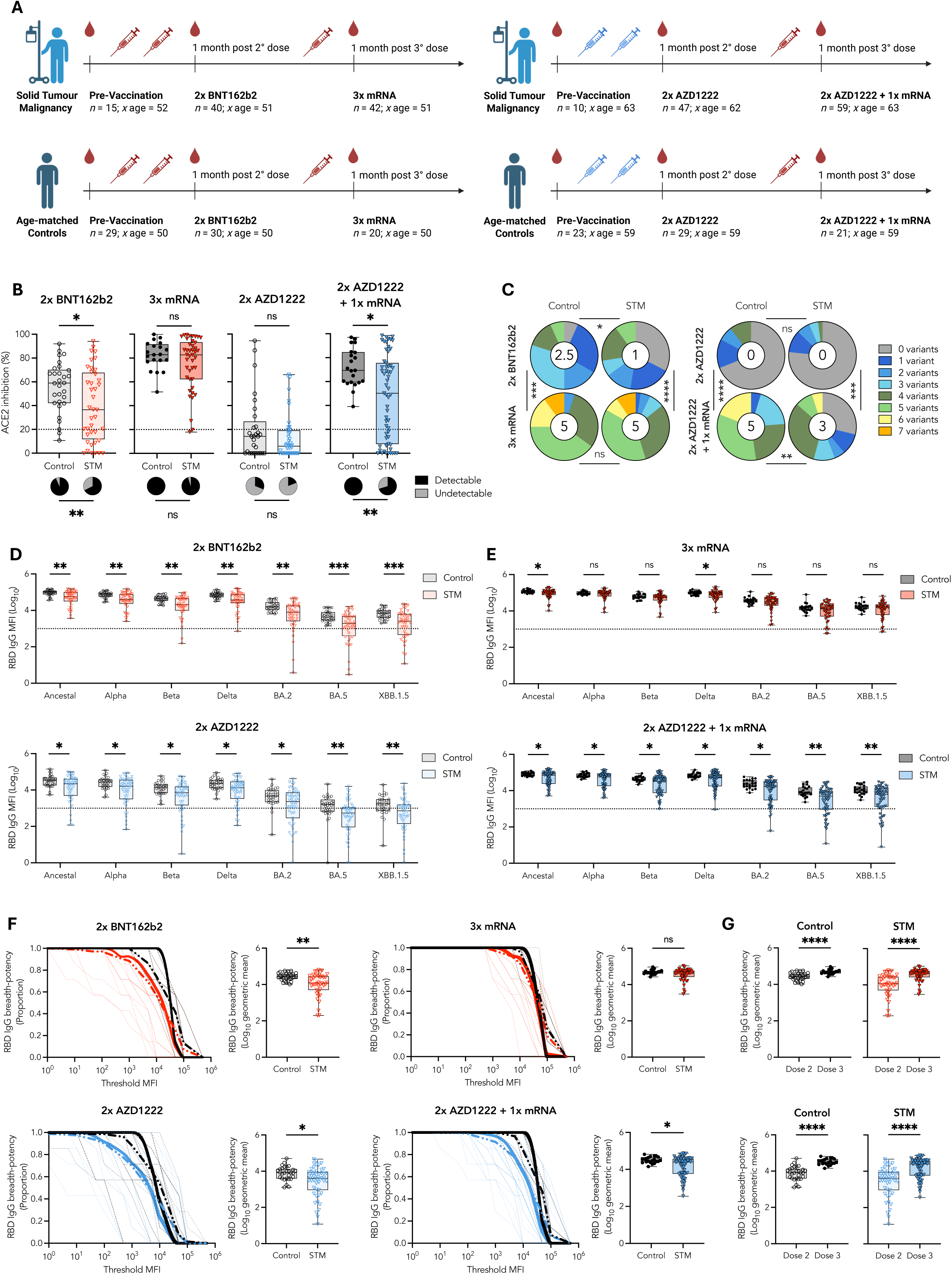
Three mRNA vaccine doses overcome impaired SARS-CoV-2 neutralization breadth-potency in STM patients. **A)** Overview of cohort demographics. **B)** Surrogate neutralization (% ACE2 binding inhibition) of ancestral SARS-CoV-2 RBD one month post second BNT162b2 or AZD1222 dose and post mRNA booster. Pie charts indicate the proportion of vaccinees for whom neutralization was (black) or was not (grey) detected. ACE2 inhibition above 20% was deemed detectable neutralization capacity, as described in methods. **C)** Proportion of vaccinees for whom neutralization was detected for the indicated number of variants. Number in the center of each pie chart indicates the median number of variants neutralized. RBD-specific IgG responses across the panel of 7 tested SARS-CoV-2 variants one month post **D)** two or **E)** three doses of the indicated vaccine regimen. **F)** RBD-specific IgG breadth-potency curves and scores one-month post indicated vaccine regimen. The proportion of variant-specific responses above a given threshold MFI is indicated by thin dashed lines for individual vaccinees. Group-specific mean proportions are indicated by thick dashed lines. Thick solid lines (breadth-potency curves) indicate the proportion vaccinees for whom the geometric mean response across variants is at or above a given threshold MFI. **G)** RBD-specific IgG breadth-potency scores one-month post dose two and dose three for BNT162b2-and AZD1222-primed vaccinees. Black: Controls; Red: BNT162b2-primed STM; Blue: AZD1222-primed STM. Box plots represent median and IQR. Horizontal dotted lines indicate positivity threshold, calculated as described in methods. Statistical differences in proportions of vaccinees with **B)** detectable surrogate neutralization and **C)** number of variants neutralized determined via chi-square test. Mann-Whitney *U*-tests performed between STM and control vaccinees within each vaccinee regimen. *p* < 0.0001 (****); *p* < 0.001 (***); *p* < 0.01 (**); *p* < 0.05 (*); non-significant (ns). MFI: median fluorescence intensity.

Assessment of SARS-CoV-2 neutralization capacity via an ACE2 inhibition assay identified significantly reduced neutralizing capacity in STM patients, as compared to age-matched controls, one month post second BNT162b2 mRNA vaccination (Fig. 1B). Both the magnitude of responses (38% reduction; *p* = 0.028) and proportion of individuals with detectable neutralizing capacity (67.5% vs 93.3%; *p* = 0.009) were significantly lower in STM compared to control vaccinees. Encouragingly, a third mRNA vaccine restored neutralizing capacity to that measured for age-matched controls (Fig. 1B). In contrast, primary AZD1222 vaccination failed to induce detectable neutralizing antibodies in the majority of STM patients and controls (80.9% and 69.0%, respectively). Although an mRNA booster vaccine significantly increased neutralizing capacity in AZD1222-primed STM patients and controls (Supplementary Fig. 1A), STM patients still generated significantly lower neutralizing responses in terms of both magnitude (28% reduction; *p* = 0.015) and proportion (70.2% vs 100%; *p* = 0.004) of responders (Fig. 1B).

Emergence of successive SARS-CoV-2 variants of concern (VoCs) have driven serial escape from neutralizing responses. We further explored the breadth of the neutralizing response against SARS-CoV-2 VoC RBDs. Neutralizing breadth induced by two mRNA vaccines was significantly lower in STM compared to control vaccinees (median 1 and 2.5 of 7 tested strains, respectively; *p* = 0.044). A third mRNA booster dose significantly increased neutralizing breadth to a median 5 strains in both STM and control vaccinees (Fig. 1C) whereas AZD1222-primed, mRNA-boosted STM patients neutralized significantly fewer variants compared to controls (median 3 and 5, respectively; *p* = 0.004) (Fig. 1C).

Given the enhanced sensitivity of IgG assessment and robust correlation of RBD-specific IgG with ACE2 inhibition (Supplementary Fig. 1B), we further examined IgG responses to VoC RBDs. Across the panel of variants, RBD-specific IgG responses were significantly lower in STM patients compared to controls following two BNT162b2 or two AZD1222 doses (Fig. 1D). An mRNA booster vaccine was required to generate appreciable IgG cross-reactivity against BA.2, BA.5, and XBB.1.5 variant RBDs in both STM patients and control vaccinees, regardless of priming vaccine regimen. However, ∼25% of STM vaccinees who received an AZD1222 prime plus mRNA booster regimen still did not generate detectable BA.5 nor XXB1.5 RBD cross-reactive IgG responses (Fig. 1E). The cumulative magnitude of cross-reactive RBD-specific IgG responses was quantified as breadth-potency scores describing the geometric mean of IgG responses across variants (Fig. 1F). Mirroring surrogate neutralization breadth data, RBD IgG breadth-potency was significantly lower in STM compared to control vaccinees following two vaccine doses, regardless of regimen (Fig. 1F). A third mRNA vaccine significantly increased cross-reactive RBD IgG responses (Fig. 1G) such that RBD IgG breadth-potency became comparable in STM patients and controls who received three mRNA doses (Fig. 1F). In contrast, the difference in RBD IgG breadth-potency between AZD1222-primed STM and control vaccinees following two AZD1222 doses was not resolved by an mRNA booster dose (*p* = 0.016) (Fig. 1F), despite significant increases in breadth-potency for both patients and controls (*p* < 0.0001) (Fig. 1G). These data highlight the value of mRNA-versus adenoviral vector-based vaccination strategies for induction of broad and potent neutralizing responses in immunocompromised groups.

Memory B cells preserve the diversity of the germinal center (GC) output and are thus critical for recall of cross-reactive antibody responses upon SARS-CoV-2 variant challenge^33^. Importantly, in a subset of vaccinees for whom peripheral blood mononuclear cells (PBMCs) were available (*n* = 10 control; *n* = 17 STM), we observed no significant differences between STM and controls in the frequency of switched CD19^+^IgD^-^ (Supplementary Fig. 2A, B) nor CD19^+^IgG^+^ (Supplementary Figure 2C, D) RBD-specific memory B cells one month post dose 2 mRNA vaccination, in line with previous characterizations of vaccine-specific memory B cell responses in other high-risk groups^3, 4, 34^. RBD-specific switched CD19^+^ B cells did not correlate with RBD-specific IgG levels (Supplementary Fig. 2B, D), as expected^3, 35^.

### Two BNT162b2 doses induce robust Fc effector functions in STM patients

We next assessed ancestral SARS-CoV-2 spike trimer-specific antibody-dependent cellular phagocytosis (ADCP) and cytotoxicity (ADCC) in STM and control vaccinees. Two BNT162b2 doses induced comparable trimer-specific ADCP and ADCC in STM patients and controls (Fig. 2A, B). In contrast, two AZD1222 doses induced significantly lower magnitude ADCP and ADCC responses in STM patients (*p* = 0.015 and *p* < 0.0001, respectively), and ADCP was induced in a lower proportion of STM patients than controls (*p* = 0.018) (Fig. 2A, B). ADCP and ADCC are largely mediated via activation of FcγRIIa and FcγRIIIa, respectively^28^. As expected, cell-based ADCP and ADCC responses strongly correlated with trimer-specific IgG engagement of soluble FcγRIIa and FcγRIIIa dimers, respectively (Fig. 2A, B), supporting further assessment of Fc functional capacity via multiplex quantification of IgG-FcγR binding responses at additional timepoints and across SARS-CoV-2 VoCs. Strong positive correlations between cell-based functional capacity and trimer-specific IgG engagement of FcγRs were observed for both the higher-affinity (FcγRIIaH and FcγRIIIaV) (Fig. 2A, B) and lower-affinity (FcγRIIaR and FcγRIIIaF) (Supplementary Fig. 3) FcγR polymorphic variants.

**Fig. 2.**
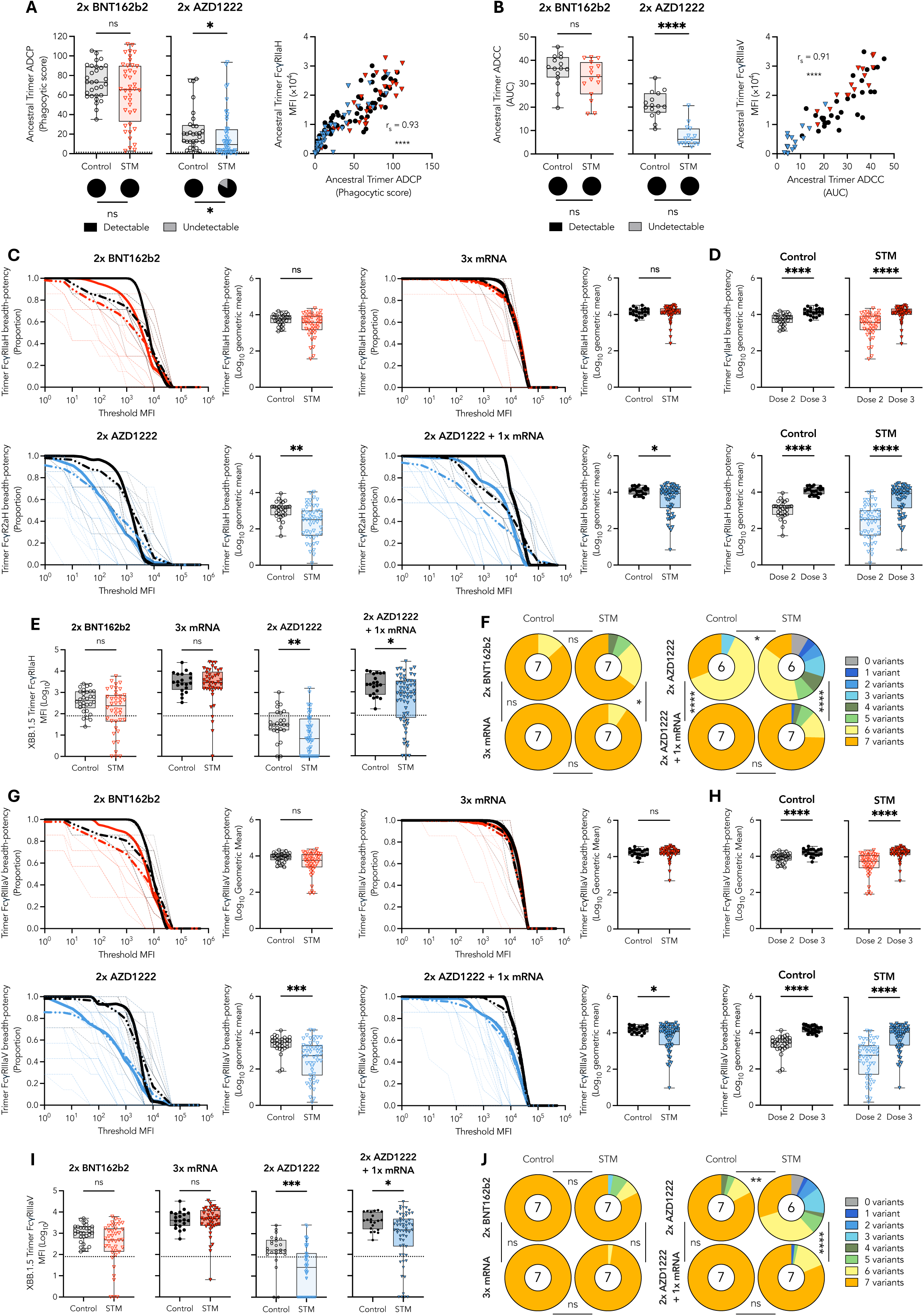
mRNA vaccination induces cross-reactive Fc effector functions in STM patients. **A)** Ancestral trimer-specific ADCP one month post two BNT162b2 or AZD1222 doses and Spearman correlation with FcγRIIaH binding. A phagocytic score above 1.4 was deemed a detectable positive response, as described in methods. **B)** Ancestral trimer-specific ADCC one month post two BNT162b2 or AZD1222 doses and Spearman correlation with FcγRIIIaV binding. An ADCC AUC above 0 was deemed a detectable positive response, as described in methods. Trimer breadth-potency curves and scores for **C)** FcγRIIaH binding or **G)** FcγRIIIaV one month post indicated vaccine regimen. The proportion of variant-specific responses above a given threshold MFI is indicated by thin dashed lines for individual vaccinees. Group-specific mean proportions are indicated by thick dashed lines. Thick solid lines (breadth-potency curves) indicate the proportion of vaccinees for whom the geometric mean response across variants is at or above a given threshold MFI. Black: Controls; Red: BNT162b2-primed STM; Blue: AZD1222-primed STM. Trimer breadth-potency scores for **D)** FcγRIIaH and **H)** FcγRIIIaV binding one month post indicated vaccine regimen. Omicron XXB.1.5 trimer-specific **E)** FcγRIIaH and **I)** FcγRIIIaV binding responses one month post indicated vaccine regimen. Proportion of vaccinees for whom **F)** FcγRIIaH and **J)** FcγRIIIaV binding was detected for the indicated number of variants. Number in the center of each pie chart indicates the median number of variants recognized. Horizontal dotted lines indicate positivity threshold. Statistical differences in proportions of vaccinees with detectable **A)** ADCP, **B)** ADCC, **F)** FcγRIIaH or **J)** FcγRIIIaV responses determined via chi-square test. Mann-Whitney *U*-tests performed between STM and control vaccinees within each vaccinee regimen. *p* < 0.0001 (****); *p* < 0.001 (***); *p* < 0.01 (**); *p* < 0.05 (*); non-significant (ns). MFI: median fluorescence intensity. AUC: area under curve.

### STM patients maintain robust Fc effector functions across variants of concern

Assessment of FcγRIIa breadth-potency (surrogate ADCP) revealed that two mRNA doses induced comparable cross-reactive responses in STM and control vaccinees, whereas STM patients who received two AZD1222 vaccines induced significantly lower FcγRIIa breadth compared to controls (*p* = 0.001) (Fig. 2C). An mRNA booster significantly enhanced FcγRIIaH responses in all groups (*p* < 0.0001) (Fig. 2D), facilitating comparable ancestral trimer-specific Fc functional capacity in STM patient and control vaccinees regardless of vaccine regimen (Supplementary Fig. 4) and comparable breadth-potency in STM and control vaccinees who received three mRNA doses (Fig. 2C). However, FcγRIIaH breadth-potency remained significantly lower in AZD1222-primed STM patients (*p* = 0.041) (Fig. 2C), driven by significantly lower magnitude responses to Omicron strains (Fig. 2E and Supplementary Fig. 4), despite both STM and control vaccinees developing FcγRIIaH responses against a median of 7 strains (Fig. 2F).

Spike trimer-specific FcγRIIIa engagement (surrogate ADCC) was similarly influenced by vaccine dose and platform, with two mRNA doses inducing broad and potent Fc functional capacity in both STM and control vaccinees (Fig. 2G–J). In contrast, STM patients who received two AZD1222 vaccines developed significantly lower FcγRIIIa breadth-potency (*p* = 0.0001) and reduced cross-reactive binding capacity (median 6 strains) compared to controls (median 7 strains) (*p* = 0.001) (Fig. 2G–J). Although an mRNA booster significantly increased cross-reactive responses to be proportionally comparable between STM and controls (median 7 strains in both groups) (Fig. 2J), FcγRIIIa breadth-potency remained significantly lower in STM patients (*p* = 0.037) (Fig. 2G) owing to significantly reduced Omicron trimer cross-reactivity (Fig. 2I and Supplementary Fig. 4).

### S2-specific IgG underpins comparable Fc effector functions in STM vaccinees

The comparable trimer-specific Fc functional capacity—measured as FcγR and complement component 1q (C1q) engagement—between STM and controls following two BNT162b2 doses was underpinned by equivalent responses against the antigenically conserved S2 domain (Fig. 3A and Supplementary Fig. 5A). In contrast, three mRNA vaccinations were required for STM and control vaccinees to generate comparable FcγR and C1q responses against the more antigenically novel spike 1 (S1) and RBD domains (Fig. 3A and Supplementary Fig. 5A). Fc functional capacity against all SARS-CoV-2 spike subunits (trimer, S2, S1, and RBD) was significantly lower in STM patients compared to controls following two AZD1222 doses and an mRNA-based booster dose was required for AZD1222-primed STM patients and controls to generate comparable S2-specific responses (Fig. 3B and Supplementary Fig. 5B).

**Fig. 3.**
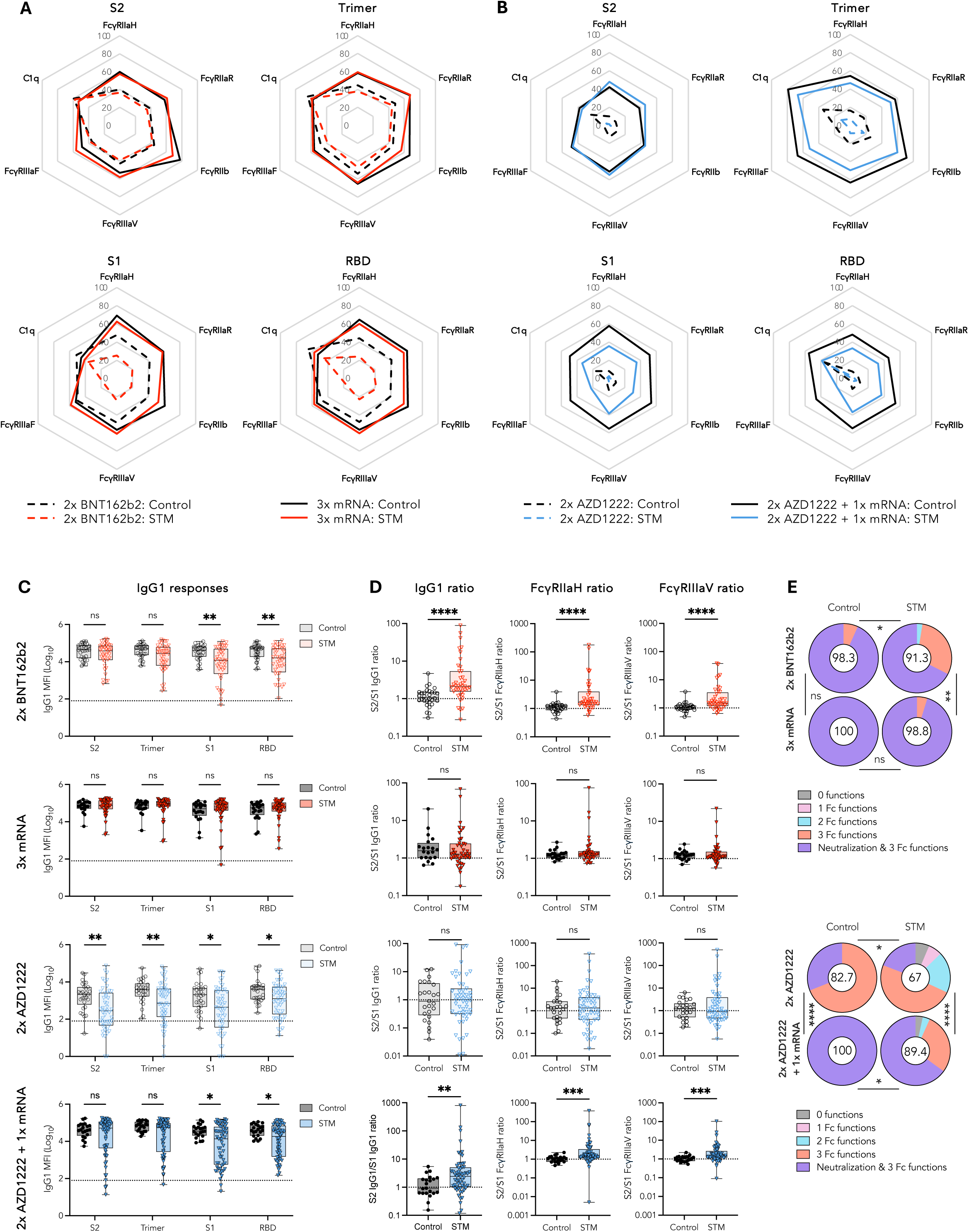
Robust vaccine-specific Fc effector functions in STM patients underpinned by a robust anti-S2 IgG response. Radar plots depicting median ancestral SARS-CoV-2 antigen-specific FcγR and C1q engagement one month post **A)** two BNT162b2 and two BNT162b2 plus mRNA booster (red) or **B)** two AZD1222 doses and two AZD1222 doses plus mRNA booster (blue). Responses normalized to maximum MFI within each Fc feature and depicted as percentiles. **C)** SARS-CoV-2 antigen-specific IgG1 responses and **D)** S2/S1 IgG1, FcγRIIaH, and FcγRIIIaV ratios one month post indicated vaccine regimen. **E)** Proportion of vaccinees for whom responses were detected for the indicated number of functions (including neutralization, FcγRIIaH, FcγRIIIaV, and C1q engagement). Number in the center of each pie chart indicates the polyfunctionality index calculated as described in methods. Box plots represent median and IQR. In **C)** horizontal dotted lines indicate positivity threshold. In **E)** statistical differences in proportions of vaccinees with detectable responses for the indicated number of functions determined via chi-square test. Mann-Whitney *U*-tests performed between STM and control vaccinees within each vaccinee regimen. *p* < 0.0001 (****); *p* < 0.001 (***); *p* < 0.01 (**); *p* < 0.05 (*); non-significant (ns). MFI: median fluorescence intensity.

Trends in spike subunit-specific FcγR engagement (Fig. 3A, B) were driven by subunit-specific IgG1—the subclass best correlated with FcγRIIa and FcγRIIIa engagement post dose 2 (Supplementary Fig. 6)^18, 36^. Equivalent spike trimer- and S2-, but significantly weaker S1- and RBD-specific, IgG1 responses were observed in two-dose mRNA vaccinated STM patients (Fig. 3C). This underpinned S2/S1 IgG1, FcγRIIa and FcγRIIIa ratios biased towards S2-specific responses (*p* < 0.0001) (Fig. 3D). Following a third mRNA vaccine, IgG1 against all SARS-CoV-2 spike subunits were comparable in STM and controls (Fig. 3C), resolving the S2-skewed S2/S1 ratios (Fig. 3D). Assessment of the functional breadth of antibody responses—neutralization, surrogate ADCP, surrogate ADCC, and complement deposition—was calculated as a weighted polyfunctionality index. STM patients who received two BNT162b2 doses generated reduced polyfunctionality compared to controls (91.3 versus 98.3; *p* = 0.032), driven by reduced neutralizing capacity which was resolved upon a booster mRNA dose (98.8 versus 100) (Fig. 3E).

In contrast, following two AZD1222 doses, IgG1 responses against all spike subunits were significantly lower in STM compared to control vaccinees (Fig. 3C) facilitating equivalent S2/S1 ratios in STM patients and control vaccinees (Fig. 3D). An mRNA booster increased IgG1 responses against the antigenically conserved S2 domain, and by extension the whole spike trimer, in both STM and control vaccinees, while S1- and RBD-specific responses remained significantly lower in STM vaccinees (Fig. 3C). This subunit bias altered S2/S1 IgG1 and FcγR ratios in STM compared to control vaccinees (*p* = 0.007 and *p* < 0.0001, respectively) (Fig. 3D), mirroring the responses in STM patients who received two BNT162b2 doses and contributing to comparable S2- and trimer-specific Fc functions in STM patients and controls (Fig. 3B). However, following an mRNA booster, polyfunctionality remained significantly lower in AZD1222-primed STM patients compared to control vaccinees (89.4 versus 100; *p* = 0.019), driven by reduced neutralizing capacity (Fig. 3E). These data suggest that two doses of mRNA vaccination can induce robust S2 antibody responses in STM patients, underpinning broadly cross-reactive Fc effector functions, equivalent to responses overserved in controls. However, STM patients require three doses of mRNA vaccination to induce robust IgG responses to novel epitopes, such as RBD, which are essential for neutralization and polyfunctionality.

### IgG subclass switching is delayed in STM patients and against variants

IgG subclass distribution heavily influences quality of the FcγR response^37, 38^, given the varied affinities of different subclasses for each FcγR^39^. Consequently, characterization of IgG subclass dynamics, as the immunoglobulin heavy constant gamma (*IGHG*) gene irreversibly switches from *IGHG3* to *IGHG4* (Fig. 4A), is critical to understanding functional antibody responses^40^.

**Fig. 4.**
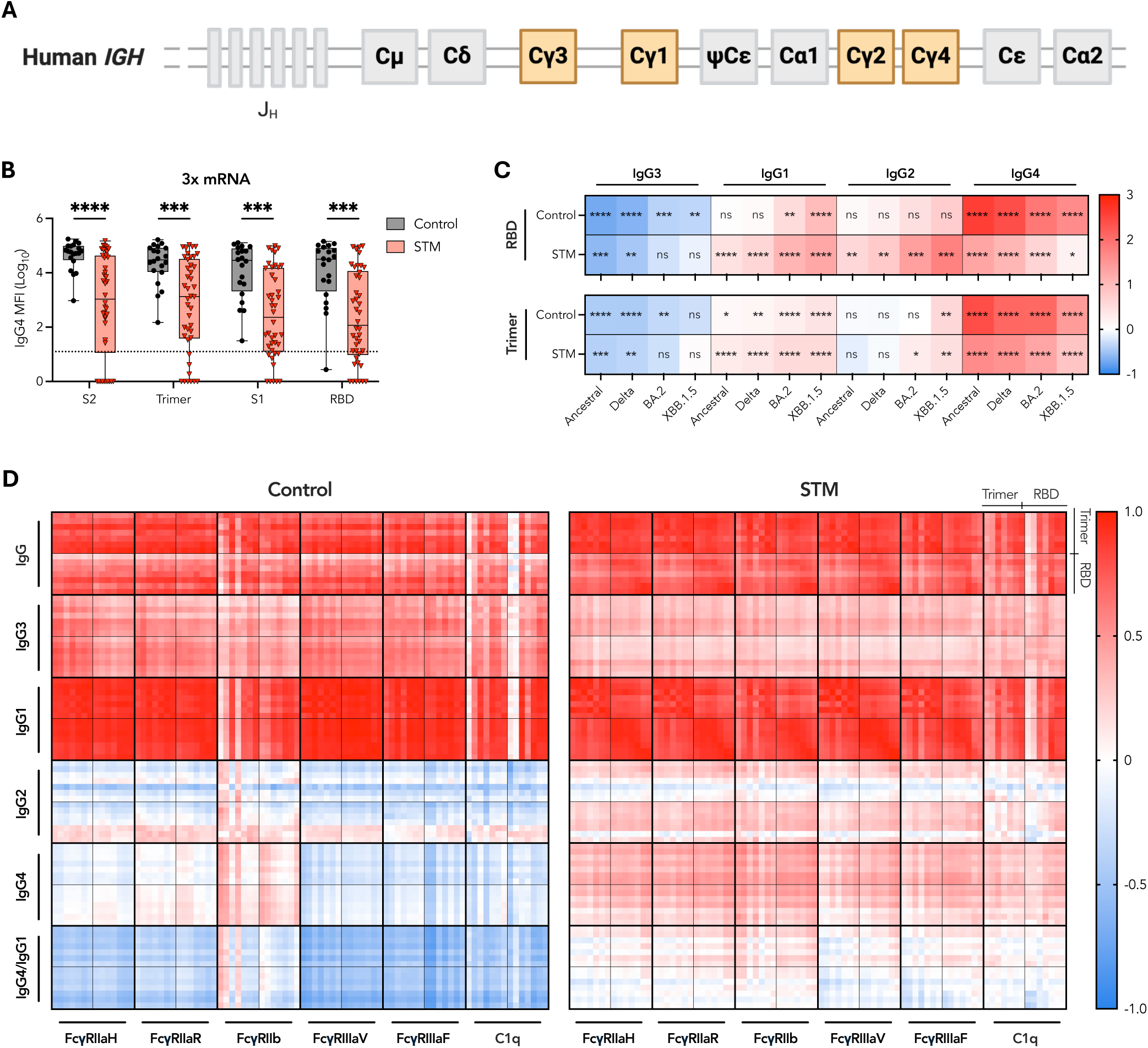
Delayed IgG subclass switching in 3x mRNA-vaccinated STM patients. **A)** Schematic of immunoglobulin heavy constant gamma (*IGHG*) gene. **B)** SARS-CoV-2 antigen-specific IgG4 responses one month post third mRNA dose. Box plots represent median and IQR. Horizontal dotted lines indicate positivity threshold. **C)** Heatmap depicting the subtracted difference in median RBD- and Trimer-antigen specific subclass responses one month post third dose minus one month post second dose for each variant. Data was z-scored prior to use. **D)** Heatmap depicting Spearman correlation coefficients describing association of variant (ancestral, alpha, beta, delta, BA.2, BA.5, XBB.1.5) trimer and RBD IgG, IgG1-4 subclass responses, and IgG4/IgG1 ratios with engagement of the indicated FcγR or C1q. Mann-Whitney *U*-tests performed between STM and control vaccinees or between dose 2 and dose 3 responses within each vaccinee regimen. *p* < 0.0001 (****); *p* < 0.001 (***); *p* < 0.01 (**); *p* < 0.05 (*); non-significant (ns). MFI: median fluorescence intensity.

Profiling of IgG subclass responses to primary BNT162b2 or AZD1222 adenoviral vector vaccination plus mRNA booster revealed distinct modulation of IgG subclass switching influenced by malignancy, vaccine platform, and SARS-CoV-2 viral variant. AZD1222-primed, mRNA-boosted STM patients and control vaccinees developed an IgG1 and IgG3 dominated response to SARS-CoV-2 antigens one month post prime and post boost vaccination (Supplementary Fig. 7)^41, 42^. In contrast, one month post third mRNA dose, the SARS-CoV-2-specific IgG subclass distribution in control vaccinees shifted from an IgG1 and IgG3 dominated response generated post second dose to an IgG1 and IgG4 dominated response, as previously reported (Supplementary Fig. 7)^38, 43, 44^. However, in STM patients, this pattern of subclass switching to IgG4 was markedly reduced (*p* < 0.001) (Fig. 4B).

Interestingly, the delayed IgG3 to IgG4 subclass switching dynamics observed in STM patients closely mirrored the delayed subclass switching against SARS-CoV-2 variants in controls (Fig. 4C). With progressively increased antigenic distance from ancestral RBD, progressively smaller fold decreases in IgG3, larger fold increases in IgG1 and IgG2, and smaller fold increases in IgG4 levels were observed (Fig. 4C). This variant-specific delay in SARS-CoV-2 IgG subclass switching was amplified in STM patients, with fold changes in variant-specific subclass levels further reduced from those calculated for controls (Fig. 4C). Since total IgG titers against variants are increasingly diminished with increased antigenic distance from ancestral SARS-CoV-2 (Fig. 1E–1F), these data suggest an overall IgG titer-dependent regulatory mechanism of subclass switching.

Elevated IgG4 in controls drove significant negative correlations of IgG4 and IgG4/IgG1 ratios against FcγR, particularly FcγRIIIaV and FcγRIIIaF, engagement following three mRNA doses (Fig. 4D), as previously described^38, 44^. However, negative correlations were conspicuously absent in STM patients (Fig. 4D), further highlighting the resilience of Fc functions in immunocompromised patients experiencing inflammatory comorbidities.

FcγR-IgG engagement is further modulated by Fc glycosylation of antigen-specific IgG at asparagine 297^45, 46^. Lower fucosylation facilitates enhanced FcγRIIIa binding and improved ADCC^45^, whereas higher galactosylation can be associated with enhanced FcγRIIa engagement and ADCP^46^. One month following two BNT162b2 doses, RBD-specific IgG1 was similarly glycosylated in STM patients and controls (Supplementary Fig. 8A). In contrast, RBD-specific IgG1 was significantly less galactosylated, but equivalently fucosylated, in STM patients compared to controls who received two AZD1222 doses (Supplementary Fig. 8B). Reduced IgG1 galactosylation may have contributed to reduced ADCP in AZD1222-primed STM patients (Fig. 2A), as suggested by the positive correlation of ADCP with galactose abundance (Supplementary Fig. 8B)^46^. However, IgG1 titers remained the strongest driver of Fc functional capacity (Supplementary Fig. 6).

### Reduced neutralizing antibodies and IgG against novel epitopes are associated with dysregulated inflammation

To begin to probe potential mechanisms underlying impaired SARS-CoV-2 neutralizing capacity despite robust Fc functions in STM patients who received two BNT162b2 doses, we assessed whether a range of inflammatory biomarkers may be associated with impaired antibody responses. Decreased galactosylation of bulk IgG Fc N-linked glycans is associated with increased age^47^ and presence of comorbidities^3^ (Fig. 5A). Expanded IgD^-^CD27^-^ double negative (DN) B cell populations (alternatively referred to as age-associated or atypical B cells) (Figure 5B) are associated with increased age^34^ and have recently been associated with STM^48^. Elevated inflammatory cytokine concentrations are also associated with STM and an inflammaging phenotype and may modulate a network of inflammatory dysregulation^31, 32^.

**Fig. 5.**
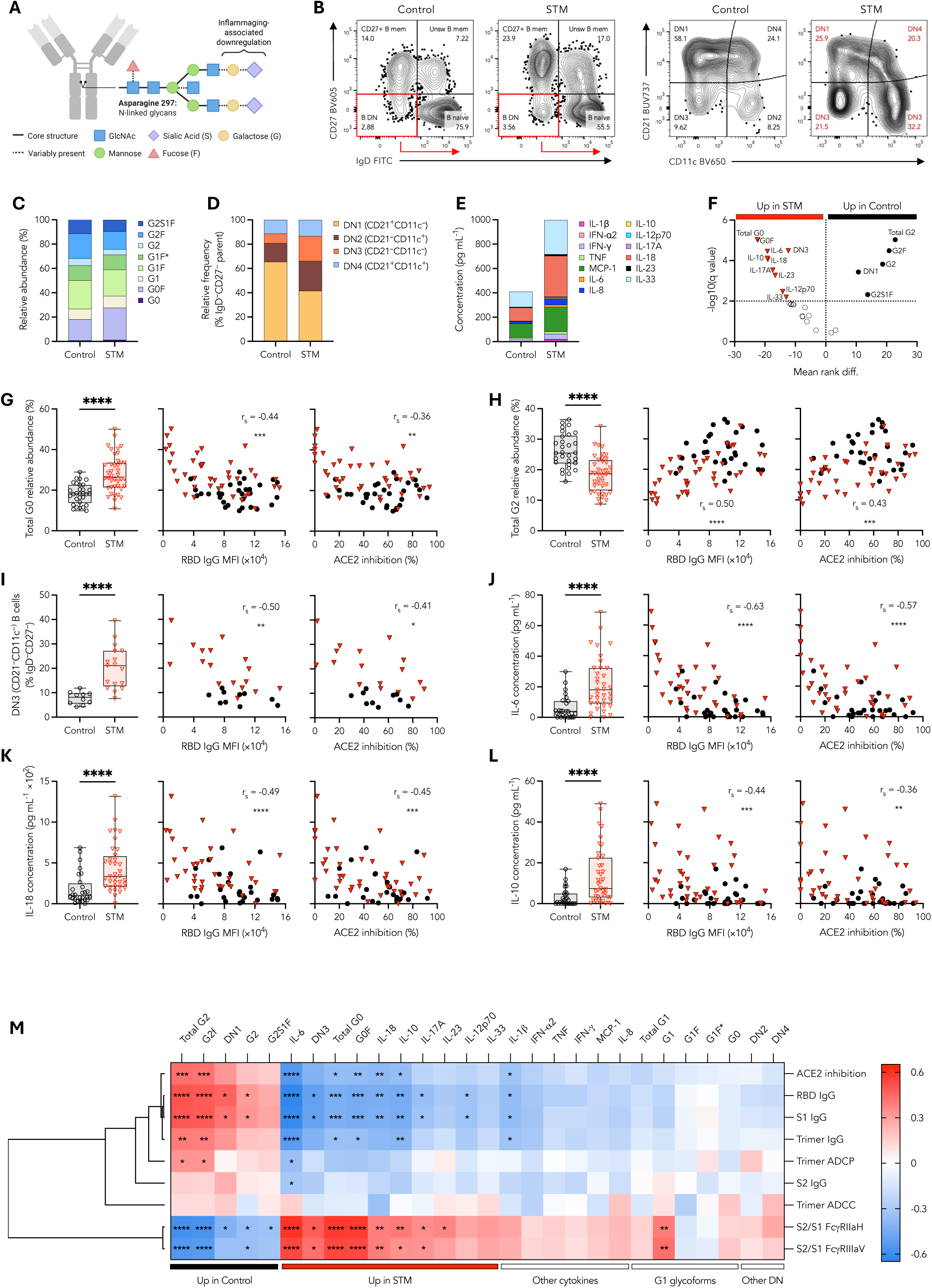
Elevated inflammatory biomarkers and DN3 B cells in STM patients correlate with impaired SARS-CoV-2 neutralizing capacity and RBD IgG. **A)** Schematic of N-linked glycans present on IgG Fc Asparagine 297**. B)** Representative FACS plots depicting identification of IgD^-^CD27^-^ double negative (DN) memory B cells. Stacked bar graphs depicting **C)** relative abundance of IgG glycoforms, **D)** relative frequencies of DN B cell populations, and **E)** median cytokine concentrations. **F)** Volcano plot of IgG glycoforms, DN B cells, and cytokine biomarkers. Relative abundance of **G)** Total G0, **H)** Total G2, **I)** Relative frequency of DN3 B cells and concentration of **J)** IL-6, **K)** IL-18, and **L)** IL-10 in controls or STM patients one month post two BNT162b2 doses. Spearman correlations for each respective inflammatory biomarker with RBD-specific IgG and % ACE2 binding inhibition. **M)** Heatmap of Spearman correlation coefficients describing the association of the indicated inflammatory biomarkers with ancestral SARS-CoV-2 IgG levels and functional antibody responses. Dendrogram generated via one minus Spearman rank correlation hierarchical clustering method. Statistical significance adjusted for multiple comparisons via Holm-Šídák method. Box plots represent the median and IQR. Mann-Whitney *U*-tests performed between STM and control vaccinees within each vaccinee regimen. *p* < 0.0001 (****); *p* < 0.001 (***); *p* < 0.01 (**); *p* < 0.05 (*); non-significant (ns). MFI: median fluorescence intensity.

We observed distinct patterns in bulk IgG glycan abundance (Fig. 5C), DN B cell frequency (Fig. 5D), and cytokine concentrations (Fig. 5E) associated with STM patients compared to controls (Fig. 5F). Inflammatory G0 glycoforms and DN3 B cells were positively correlated with each other and with IL-6, IL-12-p70, IL-17A, IL-10, while G2 glycoforms and DN1 B cells were positively associated but inversely correlated with inflammatory cytokines and glycoforms (Supplementary Fig. 9), suggesting an inflammatory network of interacting cytokine, glycan, and DN B cell biomarkers. Importantly, for the subset of individuals for whom longitudinal plasma samples were available, the relative abundance of bulk IgG glycoforms was unchanged over time (Supplementary Fig. 10A), demonstrating limited influence of COVID-19 vaccination upon bulk IgG glycosylation. This suggests that levels of these inflammatory biomarkers one month post second dose vaccination are likely representative of pre-vaccination levels, as previously described^3, 49, 50^.

Relative abundance of inflammatory total agalactosylated IgG (Total G0)—calculated as the sum of both fucosylated (G0F) and afucosylated (G0) glycoforms—was increased in STM patients compared to controls (*p* < 0.0001) (Fig. 5G). Concomitantly, increased relative abundance of digalactosylated IgG (Total G2)—calculated as the sum of fucosylated (G2F), sialylated fucosylated (G2S1F), and afucosylated (G2) glycoforms containing two galactose units—was observed in controls (*p* < 0.0001) (Fig. 5H). There was no difference in total fucosylated IgG between STM patients and controls (Supplementary Fig. 10B). In STM patients, as compared to controls, we observed increased frequencies of CD11c^-^CD21^-^ DN3 B cells (*p* < 0.0001) (Fig. 5I) concomitant with decreased frequencies of CD11c^-^CD21^+^ DN1 B cells as well as increased CD11c^+^CD21^-^ DN2 B cells (Supplementary Fig. 10C). We further observed significantly increased concentrations of IL-6 (*p* < 0.0001), IL-18 (*p* < 0.0001), IL-17A (*p* = 0.0004), IL-23 (*p* = 0.001), IL-12-p70 (*p* = 0.004), and IL-33 (*p* = 0.013) in STM patients compared to controls (Fig. 5F–K) in addition to increased IFN-γ (*p* = 0.021), TNF (*p* = 0.019), and IL-8 (*p* = 0.025) in STM patients (Supplementary Fig. 10D). Immunosuppressive IL-10 was also significantly elevated in STM patients compared to controls (*p* < 0.0001) (Fig. 5F, L), likely in response to the systemic inflammatory dysregulation experienced by malignancy patients^31, 32^. The significantly elevated inflammatory IgG glycoforms, DN B cells, and cytokines associated with inflammatory dysregulation, including G0, DN3 B cells, IL-6, IL-10, IL-17A, and IL-18, were significantly negatively correlated with both ancestral RBD-specific IgG levels and neutralizing capacity (Fig. 5G–M), whereas Total G2, was significantly positively correlated with ancestral RBD-specific IgG levels and neutralizing capacity (Fig. 5H–M).

In contrast, correlations between whole spike trimer-specific ADCP or ADCC and inflammatory biomarkers were limited (Fig. 5M), underpinned by the weak or absent correlations between inflammatory biomarkers and S2-specific IgG despite the negative correlations with S1-specific IgG (Fig. 5M). We further explored whether inflammatory dysregulation may influence mRNA vaccine-induced Fc functional capacity via modulation of epitope bias. Significant positive correlations were observed for S2/S1 FcγRIIa and FcγRIIIa ratios against inflammatory biomarkers, most notably G0 glycoforms, IL-6, IL-18, and DN3 B cells as well as immunosuppressive IL-10 (Fig. 5M), whereas inverse correlations were observed with total G2 glycan species and DN1 B cells (Fig. 5M). These data suggest that while systemic inflammation may impair the development of antibody responses against novel epitopes, including RBD-specific IgG required for efficient SARS-CoV-2 neutralization, Fc effector functions targeting antigenically conserved epitopes may still be successfully generated under conditions of inflammatory dysregulation. Indeed, hierarchical clustering grouped the correlations of inflammatory biomarkers with S1- and RBD-driven responses in a distinct node, separate from S2- and trimer-driven Fc functions. Overall, these glycan, cytokine, and B cell data point to a signature of inflammatory dysregulation in STM patients that impairs development of antibody responses against novel epitopes resulting in decreased SARS-CoV-2 neutralizing capacity.

Importantly, the correlation of elevated inflammatory biomarkers with reduced vaccine-specific IgG in STM patients was absent in controls (Supplementary Fig. 11A, B), given the robust antibody responses and generally limited inflammation in this group. Inflammatory networks were limited to positive correlations between cytokines and inverse correlations between G0 and G2 glycan species within controls (Supplementary Fig. 11A). In STM patients alone, elevated IL-6 was associated with increased immunosuppressive IL-10 and DN3 B cell frequency was positively correlated with inflammatory G0 glycan species, all of which negatively correlated with reduced RBD-, S1-, and trimer-, but not S2-, specific IgG (Supplementary Fig. 11B) one month post second mRNA dose. Upon receipt of a third mRNA vaccination and resolution of differences in antibody titers between STM and control vaccinees, RBD-specific IgG and neutralization no longer correlated with inflammatory biomarkers (Supplementary Fig. 12). These data highlight the value of booster vaccine doses for individuals with chronic inflammatory comorbidities.

## DISCUSSION

Our study compared polyfunctional antibody responses induced by two-dose BNT162b2 or AZD1222 vaccination plus mRNA booster in STM patients and age-matched controls. Here, we link impaired SARS-CoV-2 neutralizing capacity and reduced RBD-specific IgG in STM patients with increased inflammatory biomarkers and an expanded CD11c^-^CD21^-^ DN3 B cell population.

Impaired neutralizing capacity was observed in STM patients compared to controls who received two BNT162b2 doses or two AZD1222 doses plus mRNA booster. Nevertheless, at these timepoints, STM patients were able to generate Fc functional capacity of comparable magnitude to that developed by control vaccinees. Importantly, both STM patients and controls induced Fc functional antibodies with preserved cross-reactivity to SARS-CoV-2 VoCs. In other immunocompromised populations, the preservation of vaccine-induced T cells, typically targeting antigenically conserved epitopes, has been described^3, 4^. Similarly, our study demonstrates the resilience of SARS-CoV-2 variant cross-reactive Fc effector functions under conditions of inflammatory dysregulation associated with impaired neutralizing responses. These data underscore the value of vaccine-induced Fc effector functions in high-risk groups^10^ and suggest a potential additional mechanism of protection against rapidly evolving viruses such as SARS-CoV-2.

Choice of vaccine platform remains an important consideration for enhanced protection in immunocompromised individuals. We found that a minimum of three mRNA vaccine doses—but not adenoviral vector-based primary vaccination plus mRNA booster—was required for STM patients to develop a neutralizing antibody response of comparable magnitude and breadth to that developed by controls. These data are consistent with prior studies of COVID-19 vaccine immunogenicity in other high-risk groups^1, 34, 51^ and emphasize the importance of booster vaccinations for immunocompromised individuals^52^. We have previously demonstrated that repeated mRNA vaccination induces elevated SARS-CoV-2-specific IgG4 antibodies that can inhibit FcγRIIIa binding and downstream Fc effector functions^38^. Intriguingly, we observed that following three mRNA vaccines STM patients generated significantly lower SARS-CoV-2-specific IgG4, while maintaining SARS-CoV-2-specific IgG1 levels comparable to controls, potentially contributing to the robust Fc functions in STM patients.

Our data suggest that differential regulation of antibody responses against antigenically conserved versus novel SARS-CoV-2 epitopes in STM compared to control vaccinees may contribute to the maintenance of Fc functions despite impaired neutralizing capacity in STM patients. Controls generated robust RBD- and S1-specific antibodies. In contrast, STM patients developed reduced RBD- and S1-specific IgG, despite IgG responses against the antigenically conserved S2 epitope being equivalent in STM and control vaccinees. These data demonstrate that reduced RBD-specific IgG in STM vaccinees is not a simple consequence of overall dampening of the prototypical response elicited in control vaccinees. Responses against antigenically conserved epitopes are robustly induced while only IgG against novel epitopes is impaired. We hypothesize that an immunosenescence-like phenotype induced by the chronic inflammatory dysregulation associated with malignancy^31,32^ may suppress GC-dependent *de novo* antibody responses against antigenically novel epitopes, underpinning impaired SARS-CoV-2 neutralization.

Reduced RBD-specific IgG and neutralizing capacity in STM patients were significantly associated with elevated inflammatory biomarkers, most notably IL-6, IL-18, G0 glycoforms and CD11c^-^CD21^-^ DN3 memory B cells. These cytokines, IgG glycoforms, and B cells were correlated in an inflammatory network, suggesting a rewired inflammatory axis in STM patients that underpins impaired vaccine responses to novel epitopes. We have previously reported that inflammatory G0 and IL-18 negatively correlate with RBD-specific IgG following two BNT162b2 doses in vaccinees with chronic inflammatory comorbidities such as renal disease and diabetes^3^. Elevated G0 has also been associated with impaired antibody responses following seasonal influenza vaccination^50^. Increased circulating CD11c^+^FcRL5^+^ atypical B cells in the elderly^34^ and increased CD11c^+^CD21^-^ DN2 and CD11c^-^CD21^-^ DN3 B cells in systemic lupus erythematosus patients^53^ have been associated with impaired SARS-CoV-2 neutralizing antibodies following two-dose AZD1222 plus mRNA booster vaccination^34^ or two-dose mRNA vaccination^53^. The repeated association of these inflammatory biomarkers with impaired antibody responses to vaccination across diverse cohorts, different vaccination regimens, and both influenza and COVID-19 vaccination suggests common mechanisms that may be universally targeted to improve response in a variety of high-risk groups.

Recently, accumulation of circulating CD11c^-^CD21^-^ DN3 B cells has been reported in STM patients^48^. These cells were hyporesponsive to B cell receptor stimulation, displaying reduced antibody production and failure to differentiate into antibody secreting cells^48^. Both DN2 and DN3 memory B cells have extrafollicular origins, and the expansion of these DN populations suggests a bias towards extrafollicular differentiation at the expense of GC formation in STM and SLE patients^48, 53^. As such, an expanded DN3 population and potential suppression of the GC responses required to develop robust antibody responses against novel epitopes may have contributed to reduced RBD-specific IgG and neutralizing capacity following COVID-19 vaccination of STM patients.

In addition to increased inflammatory cytokines, anti-inflammatory IL-10 was also elevated in STM patients as compared to controls and was similarly correlated with impaired SARS-CoV-2 neutralizing capacity. Until recently, it was considered somewhat paradoxical that elevated inflammation could occur concurrently with immunosuppression. However, IL-10— a potent immunomodulatory cytokine that serves to counterbalance the harmful effects of chronic inflammation driven by sustained IL-6 elevation—was identified as a link between increased inflammation and damped immunity^54^. Indeed, IL-6 is required to maintain high levels of IL-10^54^. Murine studies demonstrate that IL-10 released by T_fh_ cells impairs GC responses, and that blocking IL-10 receptor in aged mice improves vaccine-induced antibody titers^54, 55^. Human influenza vaccination studies have also associated elevated IL-10 in the elderly with impaired antibody responses^56, 57^. Our data, together with prior studies, suggest an inflammaging phenotype may drive immunosenescence in individuals with inflammatory co-morbidities, such as STM, and impair development of antibody responses against novel epitopes resulting in decreased SARS-CoV-2 neutralizing capacity.

Inflammatory dysregulation, such as that experienced by STM patients^31, 32^, may modulate FcγR expression on monocytes and NK cells^58^. Future studies should examine ADCP and ADCC using autologous monocytes or NK cells, respectively, from STM patients rather than THP-1 monocytes and healthy donor primary NK cells. Chronic inflammation is associated with a dysfunctional NK cell phenotype^59, 60^, but is also associated with upregulation of FcγRI under conditions of IFN-γ-induced inflammatory stress^61, 62^. As such, potentially elevated FcγRI in STM patients may enhance ADCP, while ADCC responses may be impaired by NK cell dysfunction. Nevertheless, a bias towards induction of ADCP may contribute to protection against COVID-19 as suggested by recent animal studies^30, 63^.

Vaccine-induced ADCP was associated with viremic control following SARS-CoV-2 infection of mice, particularly against Omicron challenge^30^, and ADCP in bronchial lavage fluid has been identified as a correlate of protection in macaques^63^. Future human clinical trials are needed to confirm if Fc functions are a correlate of protection, particularly in immunocompromised patients.

Although our study provides a comprehensive analysis of polyfunctional antibody responses to COVID-19 vaccination in STM patients, there are several limitations to our study that warrant further investigation. Given the urgency with which STM patients were vaccinated upon authorization of COVID-19 vaccines, limited pre-vaccination samples were available for STM patients in this study. We were also underpowered to dissect the influence of distinct STM diagnoses and treatment regimens upon vaccine immunogenicity. Furthermore, this study was not designed to compare the longitudinal dynamics of humoral responses in STM and control vaccinees. However, identification of more rapid waning of SARS-CoV-2 antibody responses in immunocompromised patients compared to healthy individuals^4^ highlights the importance of assessing the stability of antibody responses in COVID-19-vaccinated STM patients in future studies.

Overall, our study provides detailed characterization of the influence of STM-associated inflammation upon COVID-19 vaccine-induced humoral immunity in infection-naïve BNT162b2 and AZD1222 vaccinees. Our data demonstrate the robust induction of Fc functional antibodies in vaccinees with STM and highlight the potential of targeting optimised Fc effector functions via precision vaccination strategies for immunocompromised populations.

## METHODS

### Ethics Statement

Study protocols were approved by the University of Melbourne (#20734, #21560, #21626, and #13344), ACT Health (#2023.LRE.00046 and #2021.ETH.00062), Northern Sydney Local Health District (#2021/ETH00630), Austin Health (#HREC/73256/Austin-2021), the Walter and Eliza Hall Institute (#20/08), and Melbourne Health (#HREC/63096/MH-2020, #HREC/68355/MH-2020, and RMH69108) Human Research Ethics Committees.

### Participant recruitment and sample collection

Participants were enrolled in Australia during the country’s ‘zero COVID-19’ policy in 2021 prior to widespread community transmission of SARS-CoV-2. COVID-19 vaccinee blood samples were collected pre-vaccination, one month (median 30 days) post-second BNT162b2 (Pfizer-BioNTech) or AZD1222 (AstraZeneca) vaccination and one month (median 30 days) post-booster BNT162b2 or mRNA-1273 (Moderna) vaccination as previously described^64, 65^. Participant demographics and clinical characteristics are described in Supplementary Table 1. Whole blood was collected into sodium heparin or EDTA anticoagulant coated vacutainers and subjected to Ficoll-Paque (Cytiva, Uppsala, Sweden; 17144002) separation. Plasma and PBMCs were isolated and stored at −80[°C or in liquid nitrogen, respectively. PBMCs were available only for a subset of patients and controls and used to support analysis of vaccine-specific humoral responses and characterization of the influence of systemic inflammation upon SARS-CoV-2 antibody responses.

### Surrogate SARS-CoV-2 neutralization assay

A customised Luminex multiplex assay was used to assess plasma SARS-CoV-2 neutralizing antibody responses as previously described^18, 42, 44, 66^. Briefly, plasma (1:2000 final dilution) was incubated overnight on a shaker at 4°C with ancestral and variant SARS-CoV-2 RBD (Sino Biological)-coupled magnetic carboxylated beads (Bio-Rad). Beads were then washed and incubated with 12.5 µg mL^-1^ AviTag biotinylated ACE2 (gift from Dale Godfrey, Nicholas Gherardin, and Samuel Redmond, The Peter Doherty Institute for Infection and Immunity, University of Melbourne) on a shaker for 1 hour at RT, 650 rpm. Beads were washed and incubated with 4 µg mL^-1^ Streptavidin, R-Phycoerythrin Conjugate (SAPE) (Thermo Fisher Scientific) on a shaker for 1 hour at RT then washed again and the relative inhibition of ACE2-RBD binding was assessed via xMAP INTELLIFLEX. A nominal cutoff of 20% (depicted by dotted line in figures) was set as previously described^66^. Assays were repeated in duplicate.

### SARS-CoV-2 multiplex bead-based assay

A customised Luminex multiplex assay was used to assess plasma SARS-CoV-2 antibody responses as previously described^18, 44^. Briefly, a panel of SARS-CoV-2 antigens comprising ancestral SARS-CoV-2 S2 (ACROBiosystems), ancestral and variant spike trimer, S1, and RBD (Sino Biological) were coupled to magnetic carboxylated beads (Bio-Rad). Plasma was diluted to the concentration appropriate for each analyte (Supplementary Table 2) and incubated with antigen-coupled beads overnight on a shaker at 4°C, 650 rpm. Beads were washed and then incubated on a shaker for 2 hours at RT with 1.3 µg mL^-1^ mouse anti-human IgA, IgG, IgG1, IgG2, IgG3, or IgG4 or soluble FcγR or C1q (Supplementary Table 2). Beads were washed again and incubated further with 1 µg mL^-1^ SAPE (Thermo Fisher Scientific) on a shaker for 2 hours at RT before a final wash. Plates were acquired via xMAP INTELLIFLEX and median fluorescence intensity (MFI) of each isotype/subclass detector or soluble FcγR/C1q binding was assessed. Background subtraction was performed, removing background of blank (buffer only) wells and non-specific binding to Simian Immunodeficiency Virus (SIV)-coupled BSA-blocked beads. Assays were performed in duplicate as two independent experiments. A positivity threshold (depicted by dotted line in figures) was set as the mean response of pre-vaccination plasma plus two standard deviations.

### Production of Fc**γ**R ectodomain dimers

Plasmids for the ectodomain dimers of human FcγRIIb, FcγRIIa-H131, FcγRIIa-R131, FcγRIIIa-V158, and FcγRIIIa-F158 were generated as previously described^67^ and kindly provided by Mark Hogarth and Bruce Wines. Proteins were expressed in HEK293F cells (Invitrogen) cultured in Freestyle medium following transfection of plasmids using Lipofectamine Transfection Reagent (Thermo Fisher Scientific), according to the manufacturer’s instructions. Supernatants were harvested 6-7 days post transfection by centrifugation at 4000 rpm for 30 min followed by filtration of the supernatant using 0.22 μM Steritop filter units (Merck Millipore) and purification via affinity chromatography using Ni-NTA agarose beads (QIAGEN). Proteins were then biotinylated via AviTag using BirA ligase (Avidity) followed by size exclusion chromatography.

### Flow cytometric B cell phenotyping and detection of RBD-specific memory B cells

Surface staining of B cells within cryopreserved human PBMCs was performed as previously described^3, 53, 68^. SARS-CoV-2 RBD-specific B cells were identified via APC- and PE-conjugated probes, as previously described^68^. Cells were washed, fixed with 1% formaldehyde, and acquired on a BD LSR Fortessa. Antibody clones and fluorochrome details are described in Supplementary Table 3. Gating strategies for RBD-specific memory B cells and DN B cells are outlined in Supplementary Figure 2 and Figure 5B, respectively.

### Bead-based THP-1 ADCP assay

ADCP was measured using a previously described bead-based ADCP assay^18, 28, 38^. Briefly, SARS-CoV-2 ancestral spike trimer (SinoBiological) was biotinylated and coupled to 1 μM fluorescent NeutrAvidin Fluospheres (Invitrogen) overnight at 4°C. Washed antigen-coated beads were incubated with plasma (1:1600 final dilution) for 2 hours at 37°C in a 96-well U-bottom plate before addition of THP-1 monocytes (100,000 per well) followed by a 16-hour incubation under cell culture conditions. Cells were fixed and acquired by flow cytometry on a BD LSR Fortessa with a high-throughput sampler attachment (HTS). The data were analyzed using FlowJo 10.9.0, and a phagocytic score was calculated as previously described using the formula: (% bead-positive cells × mean fluorescent intensity). Assays were repeated in duplicate. A positivity threshold (depicted by dotted line in figures) was set as the mean response of pre-vaccination plasma plus two standard deviations.

### Luciferase-based ADCC assay

ADCC was examined in a subset of vaccinees (*n* = 60; *n* = 15 per cohort) using a previously described luciferase-based ADCC assay^28, 38^. NK cells from a healthy donor were enriched and purified using the EasySep Human NK Cell Enrichment Kit (STEMCELL Technologies Inc). NK cells (20,000 per well) and Ramos S-Luc cells (5000 per well) were added to 96-well V-bottom cell culture plates and incubated with fourfold sample dilutions (1:100, 1:400, and 1:1600 final dilutions) for 4 hours at 37°C. Samples were tested in duplicate, and “no antibody” and “target cell only” controls were included. Following incubation, cells were washed and developed with britelite plus luciferase reagent (Revvity). Luminescence was read using a FLUOstar Omega microplate reader (BMG Labtech). The relative light units measured was used to calculate % ADCC with the formula: (“no antibody” − “antibody sample”)/(“target cell only”) × 100. A positivity threshold (depicted by dotted line in figures) was set as the mean response of pre-vaccination plasma plus two standard deviations.

### Antigen-specific IgG glycan profiling

Glycosylation of RBD-specific IgG was analysed via mass spectrometry following isolation of antigen-specific IgG as previously described^69^. Briefly, Nunc MaxiSorp flat-bottom 96-well plates (Thermo Fisher Scientific) were coated overnight at 4 °C with 1 µg mL^-1^ RBD. Plates were washed prior to addition of plasma diluted 1:10 with PBS-Tween 20. Plates were then incubated for 1 hour at 37 °C, 400 rpm, washed again in PBS-Tween 20, PBS, and finally 50 mM ammonium acetate. RBD-specific antibodies were then eluted with 100 mM formic acid and dried for 4 hours under vacuum. The dried samples were redissolved in a mixture of acetonitrile and 100 mM ammonium acetate (15:85) and acidified trypsin (10 ng mL^-1^ in 1 mM acetic acid) and the plate was incubated overnight at 37 °C, 300 rpm. Tryptic peptides were analysed via Reverse Phase Liquid Chromatography-Mass Spectrometry (LC-MS/MS). The LC system was equipped with an Acclaim Pepmap nano-trap column (Dionex-C18, 100 Å, 75 μm × 2 cm) and an Acclaim Pepmap RSLC analytical column (Dionex-C18, 100 Å, 75 μm × 50 cm). The eluents were 0.1% (v/v) formic acid in water and acetonitrile with 0.1% (v/v) formic acid. Tryptic peptides were injected into the trap column and a gradient elution was performed (45 mins, 2-85%). The IgG1 relevant EEQYNSTYR glycopeptides had a retention time of 18 minutes in these conditions. MS was performed on a Fusion Lumos Tribrid Mass Spectrometer (Thermo Fisher Scientific). The positive mode nano-spray voltage was set to 1.9 kV. MS2 was performed with Higher-energy collisional dissociation (HCD) set to an energy of 35%, with dynamic exclusion. A targeted inclusion list was provided for G0, G1, G2, G0F, G1F, G2F. Raw data files were analysed with MSFragger in FragPipe (MSFragger version: 4.1)^70^, and the results were analysed in Skyline (Ver 23.1.0.455). The following IgG1 glycans were observed across all samples: G0, G1, G2, G0F, G1F, G2F, G0FB, G1FB, G2S1F. Intensities were recorded as the sum of the area of the first three isotope signals of each glycopeptide. The % afucosylation was determined by Equation 1, where G0, G1, G2, G0F, G1F, G2F, G0FB, G1FB, G2S1F refer to the intensity of the relevant glycopeptide.

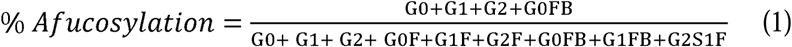

The average number of galactose units per glycan (y) was determined using Equation 2.

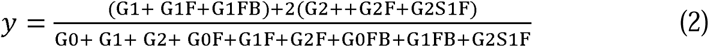

### IgG purification and IgG N-linked glycan profiling

IgG antibodies were purified from plasma and glycan species profiled as previously described^3^. Briefly, total IgG was isolated using the Melon Gel IgG Purification Kit (Thermo Fisher Scientific) according to the manufacturer’s protocol. Purified IgG samples were then centrifuged through 100 kDa Amicon Ultra Centrifugal Filters (Merck Millipore) at 14,000 *g* for 15 min to remove excess serum proteins and buffer exchange antibodies into PBS. Purity was confirmed via SDS–polyacrylamide gel electrophoresis (Bio-Rad Laboratories) and IgG concentrations were measured using a NanoDrop spectrophotometer (Bio-Rad Laboratories). IgG N-linked glycosylation patterns were measured according to the ProfilerPro glycan profiling LabChip GXII Touch protocol on the LabChip GXII Touch HT Microchip-CE platform (PerkinElmer) using the LabChip GX Touch software (v.1.9.1010.0). Microchip capillary electrophoresis laser-induced fluorescence analysis of digested and labeled N-linked glycans was performed. The relative prevalence of major N-linked glycan profiles of IgG was analyzed using the LabChip GX Reviewer (PerkinElmer) v.5.4.2222.0. Peaks were assigned based on the migration of known standards and glycan digests. The peak area and relative prevalence of each glycan pattern were calculated.

### Cytokine analysis

Donor plasma was diluted 1:2 to measure cytokines using the LEGENDplex Human Inflammation Panel 1 kit (BioLegend) according to the manufacturer’s instructions. Cytokines and chemokines including IL-1β, IFNα2, IFN-γ, TNF, MCP-1 (CCL2), IL-6, IL-8 (CXCL8), IL-10, IL-12p70, IL-17A, IL-18, IL-23 and IL-33 were measured. Samples were acquired on a FACSCanto II cytometer (BD Biosciences) and analyzed with the QOGNIT LEGENDplex online software (https://legendplex.qognit.com).

### Breadth and polyfunctionality analysis

Breadth of neutralizing or Fc responses was calculated as the number of variants (out of the seven tested) for which the response was above the calculated positivity threshold (20% inhibition or mean pre-vaccination response plus two standard deviations, respectively). The polyfunctionality index for each cohort was calculated as a weighted proportion of antibody functions (out of ACE2 inhibition, FcγRIIaH binding, FcγRIIIaV binding, and C1q binding) for which responses were detectable above the positivity threshold, calculated as (0/4)*%n_0_+(1/4)*%n_1_ +(2/4)*%n_2_+(3/4)*%n_3_+(4/4)*%n_4_, where %n_x_ is the percentage of vaccinees positive for *x* number of functions. The result is a value between 0 and 100.

### Statistical analysis

Prism GraphPad version 10.4.1 (GraphPad Software) was used to develop graphs and perform the statistical analyses described in the Figure legends. Breadth-potency scores were calculated as the geometric mean response across all seven tested variant antigens for each subject. Breadth–potency curves represent the proportion of individuals exhibiting a geometric mean antibody or Fc functional response across VoCs above a given MFI. Curves for each respective subject group were generated using the Akima spline method in Prism.

## Data Availability

The data that support the findings of this study are available from the corresponding author upon reasonable request.

## ACKNOWLEDGEMENTS

This research was funded by the National Health and Medical Research Council (NHMRC) EL2 Investigator grant #2008092 awarded to A.W.C. and NHMRC L1 Investigator grant #1173871 was awarded to K.K. A.K.W., W.S.L., T.H.O.N., K.S., S.J.K., and K.J.S. are also supported by NHMRC Investigator grants. O.H.L.W. is supported by an NHMRC Ideas grant #2029642 to S.J.R. This study was further supported by a Medical Research Future Fund (MRFF) grant #2016062 to J.A.T., A.K.W., T.H.O.N., K.K., S.J.K., and A.W.C. The COVID PROFILE study was supported by WHO Unity funds (2020/1085469-0), and WEHI philanthropic funds. The DISCOVER-HCP study was supported by the National Institute of Allergy and Infectious Diseases, National Institutes of Health, Department of Health and Human Services, Centers of Excellence for Influenza Research and Response (CEIRR) grant #HHSN272201400005C to the University of Rochester and a subcontract to KS. This work was made possible through Victorian State Government Operational Infrastructure Support and Australian Government NHMRC IRIISS. We thank E Haycroft and C Aurelia (University of Melbourne), G Gibney, and F James (Austin Health) for their outstanding technical assistance. We thank the COVID PROFILE team for their contribution to the sample collection and processing of samples as part of the COVID PROFILE cohort study. We thank the patients and volunteers who participated in this study for their generous contributions. Cartoon schematics were created in Biorender.com.

## AUTHOR CONTRIBUTIONS

R.A.P., A.M.F., Y.K., and A.W.C. conceived the study. R.A.P., M.K., and A.W.C designed the experiments. R.A.P., M.K., L.K., L.F.A., and O.H.L.W. performed and analyzed the experiments. A.K.W., W.S.L., B.D.W., P.M.H., and S.J.K. provided crucial reagents. J.-W.W., G.C., E.M.M., I.M., K.A.B., D.A.B., J.M.T., J.A.T., T.H.O.N., K.S., A.L., A.L.H., A.Y., H.R.W., S.M., K.K., A.M.F., and Y.K. coordinated cohort recruitment and clinical sample collection. W.S.L., P.R., K.B.A., and K.J.S. provided technical advice. S.J.R., S.J.K., K.J.S., K.K., and A.W.C supervised the project. R.A.P. and A.W.C. wrote the manuscript. All authors reviewed and approved the manuscript.

