## Supplementary Tables and Figures for "Dysregulated inflammation in solid tumor malignancy patients shapes polyfunctional antibody responses to COVID-19 vaccination"

**Supplementary Table 1.** Cohort demographics and clinical characteristics

| | 2x BNT162b2 | 2x AZD1222 | 3x mRNA | 2x AZD1222 + 1x mRNA |
|---|---|---|---|---|
| **Control, *n*** | 30 | 29 | 20 | 21 |
| Age, mean (range) | 50 (27-67) | 59 (29-82) | 50 (28-66) | 59 (42-82) |
| Female (%) | 18 (60%) | 16 (55%) | 13 (65%) | 12 (57%) |
| **STM, *n*** | 40 | 47 | 42 | 59 |
| Age, (mean, range) | 51 (26-76) | 62 (39-88) | 51 (23-84) | 63 (38-78) |
| Female, *n* (%) | 25 (63%) | 23 (49%) | 27 (64%) | 33 (56%) |
| Metastatic disease, *n* (%) | 22 (55%) | 19 (40%) | 24 (57%) | 32 (54%) |
| Malignancy, *n* (%) | | | | |
| Bone | 0 (0%) | 0 (0%) | 1 (2%) | 0 (0%) |
| Breast | 7 (17.5%) | 9 (19%) | 8 (19%) | 16 (27%) |
| Brain | 9 (22.5%) | 11 (23%) | 8 (19%) | 7 (12%) |
| Glioma | 9 (100%) | 9 (82%) | 7 (87.5%) | 6 (86%) |
| Meningioma | 0 (0%) | 2 (18%) | 0 (0%) | 1 (14%) |
| Other | 0 (0%) | 0 (0%) | 1 (12.5%) | 0 (0%) |
| Gastrointestinal | 7 (17.5%) | 4 (8.5%) | 10 (24%) | 10 (17%) |
| Colorectal | 6 (86%) | 2 (50%) | 6 (60%) | 3 (30%) |
| Gastric | 0 (0%) | 0 (0%) | 0 (0%) | 1 (10%) |
| GIST | 0 (0%) | 0 (0%) | 1 (10%) | 3 (30%) |
| Hepatic | 0 (0%) | 0 (0%) | 1 (10%) | 1 (10%) |
| Oesophageal | 0 (0%) | 0 (0%) | 0 (0%) | 1 (10%) |
| Pancreatic | 1 (14%) | 2 (50%) | 2 (20%) | 1 (10%) |
| Genitourinary | 7 (17.5%) | 6 (13%) | 6 (14%) | 6 (10%) |
| Prostate | 1 (14%) | 5 (83%) | 1 (17%) | 4 (67%) |
| Testicular | 1 (14%) | 0 (0%) | 1 (17%) | 0 (0%) |
| Renal | 4 (47%) | 1 (17%) | 3 (50%) | 2 (33%) |
| Urothelial | 1 (14%) | 0 (0%) | 1 (17%) | 0 (0%) |
| Gynaecological | 4 (10%) | 4 (8.5%) | 5 (12%) | 8 (14%) |
| Cervical | 1 (25%) | 0 (0%) | 1 (20%) | 0 (0%) |
| Ovarian | 3 (75%) | 1 (25%) | 4 (80%) | 3 (37.5%) |
| Uterine | 0 (0%) | 3 (75%) | 0 (0%) | 5 (62.5%) |
| Head and Neck | 2 (5%) | 3 (6%) | 1 (2%) | 2 (3%) |
| Nasopharyngeal | 2 (100%) | 0 (0%) | 1 (100%) | 0 (0%) |
| Oropharyngeal | 0 (0%) | 3 (100%) | 0 (0%) | 2 (100%) |
| Lung | 4 (10%) | 2 (4%) | 3 (7%) | 1 (2%) |
| NSCLC | 2 (50%) | 2 (100%) | 1 (33%) | 1 (100%) |
| Mesothelioma | 2 (50%) | 0 (0%) | 2 (67%) | 0 (0%) |
| Skin | 0 (0%) | 8 (17%) | 0 (0%) | 7 (12%) |
| Melanoma | 0 (0%) | 4 (50%) | 0 (0%) | 4 (57%) |
| Other | 0 (0%) | 4 (50%) | 0 (0%) | 3 (43%) |
| Treatment, *n* (%) | | | | |
| Chemotherapy | 20 (50%) | 16 (34%) | 23 (55%) | 21 (36%) |
| Immunotherapy | 7 (17.5%) | 11 (23%) | 4 (9.5%) | 9 (15%) |
| Targeted therapy | 5 (12.5%) | 6 (13%) | 11 (26%) | 12 (31%) |
| Radiotherapy | 1 (2.5%) | 1 (2%) | 0 (0%) | 0 (0%) |
| Observation following previous chemotherapy | 5 (12.5%) | 7 (15%) | 3 (7%) | 8 (14%) |
| Observation following previous immunotherapy | 2 (5%) | 1 (2%) | 1 (2%) | 1 (2%) |
| Observation following previous radiotherapy | 0 (0%) | 3 (6%) | 0 (0%) | 1 (2%) |
| Best supportive care | 0 (0%) | 2 (4%) | 0 (0%) | 0 (0%) |

Chemotherapy: any patient who received chemotherapy; Immunotherapy: any patient who received immunotherapy, but not chemotherapy; Targeted therapy: any patient who received targeted therapy, but did not receive chemotherapy nor immunotherapy; Radiotherapy: any patient who received radiotherapy, but did not receive chemotherapy, immunotherapy, nor targeted therapy. Abbreviations: STM, solid tumor malignancy; GIST, gastrointestinal stromal tumor; NSCLC, non-small cell lung cancer.

**Supplementary Table 2.** Details of biotinylated mouse anti-human anti-IgG antibody detectors and soluble FcγR and C1q reagents used in bead-based multiplex assays

| Detector | Clone | Manufacturer | Catalogue number | Plasma dilution |
|---|---|---|---|---|
| Total IgA, biotin | MT20 | MabTech | 3860-6-250 | 1:250 |
| Total IgG, biotin | MT78/145 | MabTech | 3850-6-250 | 1:3200 |
| IgG1, biotin | HP1218 | MabTech | 3851-14-250 | 1:1600 |
| IgG2, biotin | HP6200 | MabTech | 3852-6-250 | 1:250 |
| IgG3, biotin | MTG34 | MabTech | 3853-6-250 | 1:250 |
| IgG4, biotin | MTG42 | MabTech | 3854-6-250 | 1:1600 |
| FcγR2a-H131 | N/A | Produced in-house | PMID: 27385782 | 1:600 |
| FcγR2a-R131 | N/A | Produced in-house | PMID: 27385782 | 1:600 |
| FcγR2b | N/A | Produced in-house | N/A | 1:600 |
| FcγR3a-V158 | N/A | Produced in-house | PMID: 27385782 | 1:600 |
| FcγR2a-F158 | N/A | Produced in-house | PMID: 27385782 | 1:600 |
| C1q | N/A | Sigma-Aldrich | C1740-.5MG | 1:600 |

Dilutions indicate the final dilution of plasma (following addition of bead cocktail) used in multiplex assays.

**Supplementary Table 3.** Details of antibodies used for detection of memory B cells

| Detector | Clone | Fluorochrome | Manufacturer | Antibody dilution |
|---|---|---|---|---|
| IgD | Goat polyclonal | FITC | SouthernBiotech | 1:100 |
| CD20 | 2H7 | APC-C7/NIR | BioLegend | 1:100 |
| CD14 | M5E2 | BV510 | BioLegend | 1:200 |
| CD3 | OKT3 | BV510 | BioLegend | 1:400 |
| CD8a | RPA-T8 | BV510 | BioLegend | 1:1000 |
| CD16 | 3G8 | BV510 | BioLegend | 1:300 |
| CD10 | HI10a | BV510 | BioLegend | 1:500 |
| CD27 | O323 | BV605 | BioLegend | 1:100 |
| CD11c | B-ly6 | BV650 | BD Biosciences | 1:50 |
| IgG | G18-145 | BV786 | BD Biosciences | 1:50 |
| CD19 | J3-119 | PECF594 | Beckman Coulter | 1:100 |
| CD71 | CY1G4 | PE-Cy7 | BioLegend | 1:50 |
| FcRL5 | 509F6 | BUV395 | BD Biosciences | 1:50 |
| CD21 | B-ly4 | BUV737 | BD Biosciences | 1:100 |

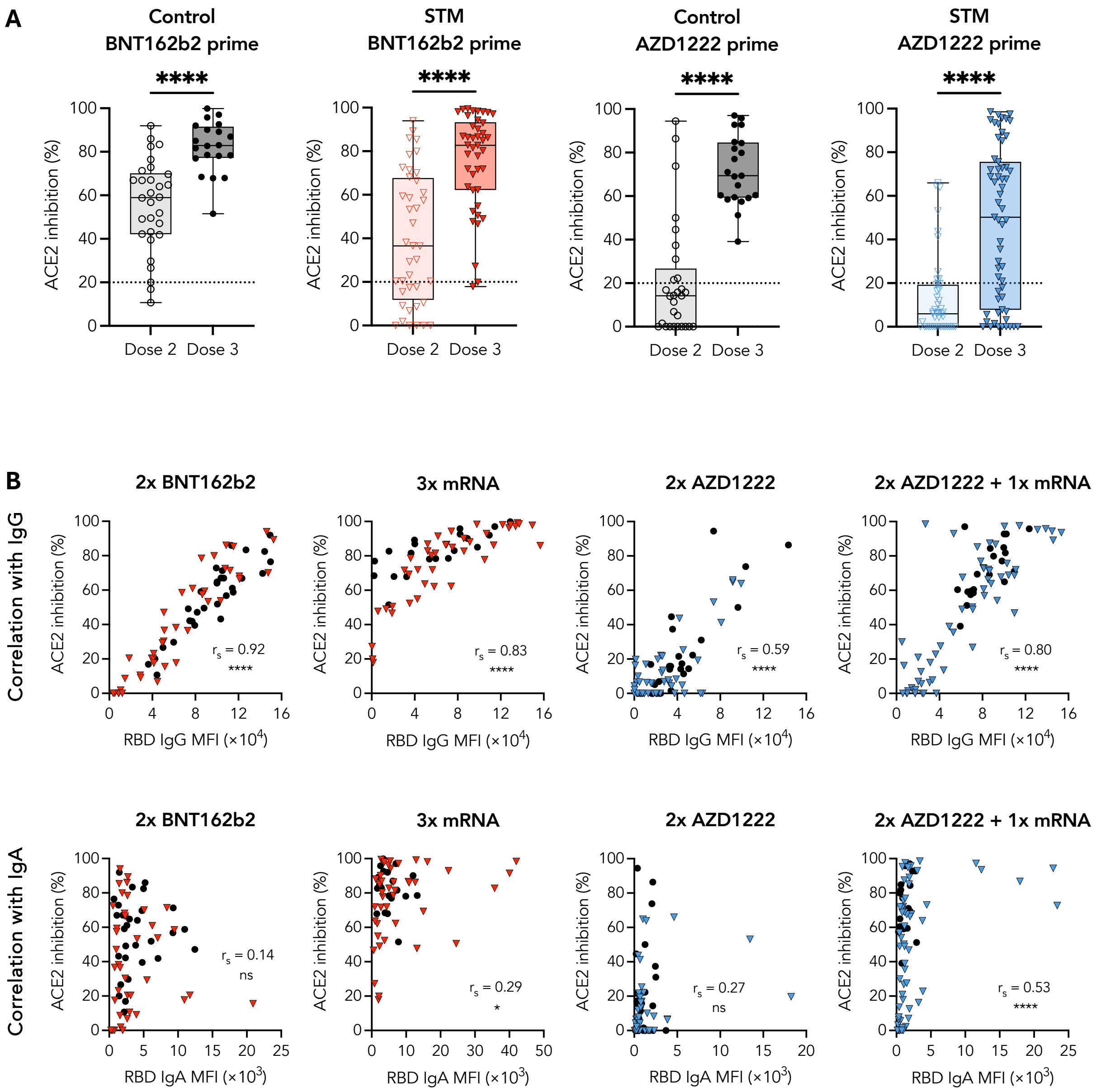


**Supplementary Fig. 1. RBD-specific IgG underpins robust neutralizing capacity following mRNA-based booster vaccination. A)** Neutralizing capacity of STM and control vaccinees one month post second (BNT162b2 or AZD1222) and one month post third (mRNA) dose of the indicated vaccine regimen. **B)** Spearman correlations of IgG or IgA with ACE2 inhibition one month post indicated vaccine regimen. Mann-Whitney *U*-tests performed between STM and control vaccinees within each vaccinee regimen. $p < 0.0001$ (****); $p < 0.05$ (*); non-significant (ns). MFI: median fluorescence intensity.

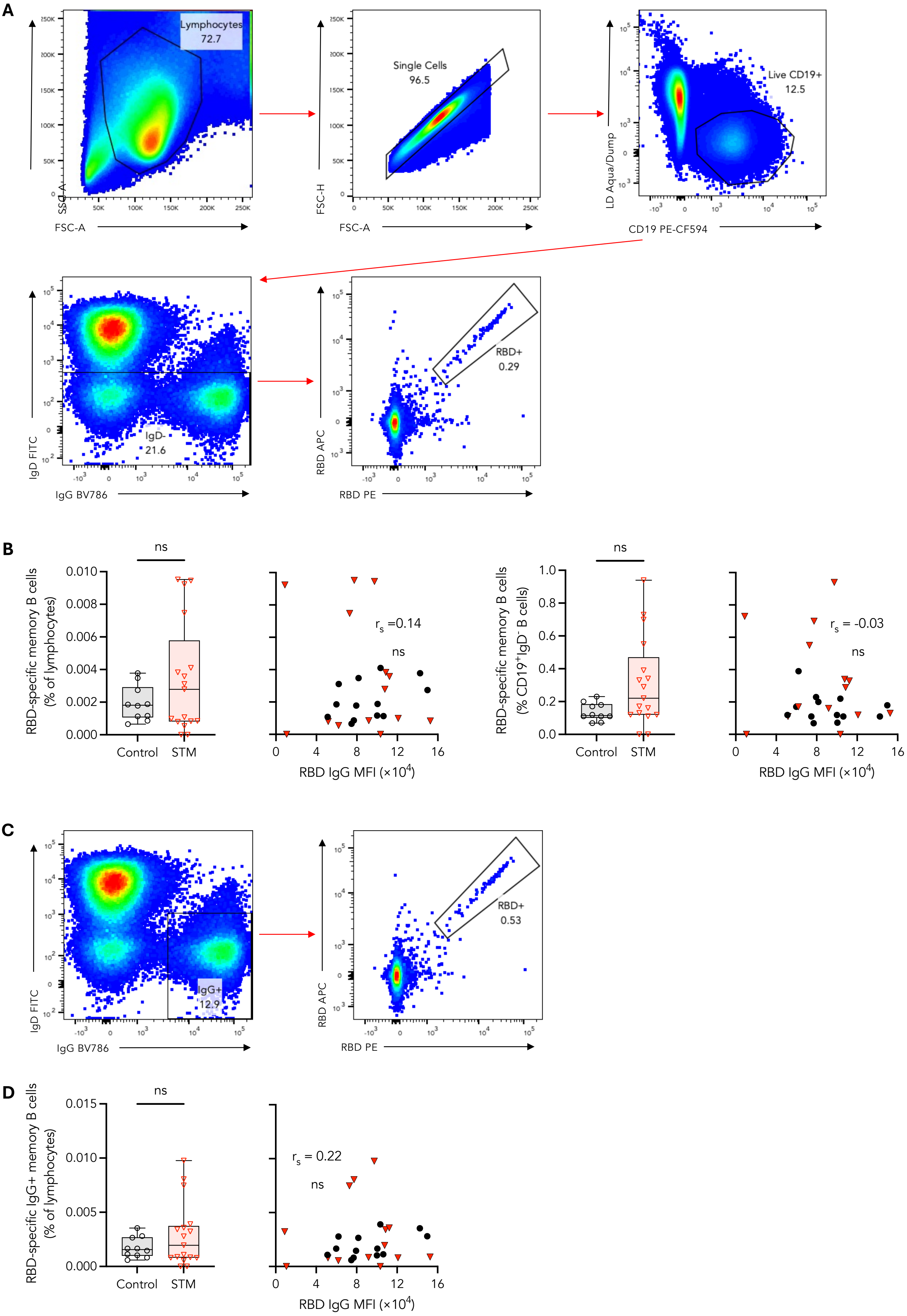
A
Lymphocytes
72.7
SSC-A
FSC-A
Single Cells
96.5
FSC-H
FSC-A
Live CD19+
12.5
LD Aqua/Dump
CD19 PE-CF594
IgD-
21.6
IgD FITC
IgG BV786
RBD+
0.29
RBD APC
RBD PE
B
ns
RBD-specific memory B cells
(% of lymphocytes)
Control
STM
r_s =0.14
ns
RBD IgG MFI (×10^4)
ns
RBD-specific memory B cells
(% CD19+IgD- B cells)
Control
STM
r_s = -0.03
ns
RBD IgG MFI (×10^4)
C
IgG+
12.9
IgD FITC
IgG BV786
RBD+
0.53
RBD APC
RBD PE
D
ns
RBD-specific IgG+ memory B cells
(% of lymphocytes)
Control
STM
r_s = 0.22
ns
RBD IgG MFI (×10^4)

**Supplementary Fig. 2. Robust RBD-specific memory B cells in STM patients. A)** Gating strategy for identification of RBD-specific switched memory B cells. **B)** RBD-specific switched memory B cells one month post second BNT162b2 dose as a frequency of $IgD^-$ B cells or total lymphocytes. Spearman correlations of RBD-specific switched memory B cells and RBD-specific IgG one month post second BNT162b2 dose. **C)** Gating strategy for identification of RBD-specific $IgG^+$ memory B cells. **D)** RBD-specific $IgG^+$ memory B cells one month post second BNT162b2 dose as a frequency of total lymphocytes. Spearman correlations of RBD-specific $IgG^+$ memory B cells and RBD-specific IgG one month post second BNT162b2 dose. Mann-Whitney *U*-tests performed between STM and control vaccinees within each vaccinee regimen. non-significant (ns). MFI: median fluorescence intensity.

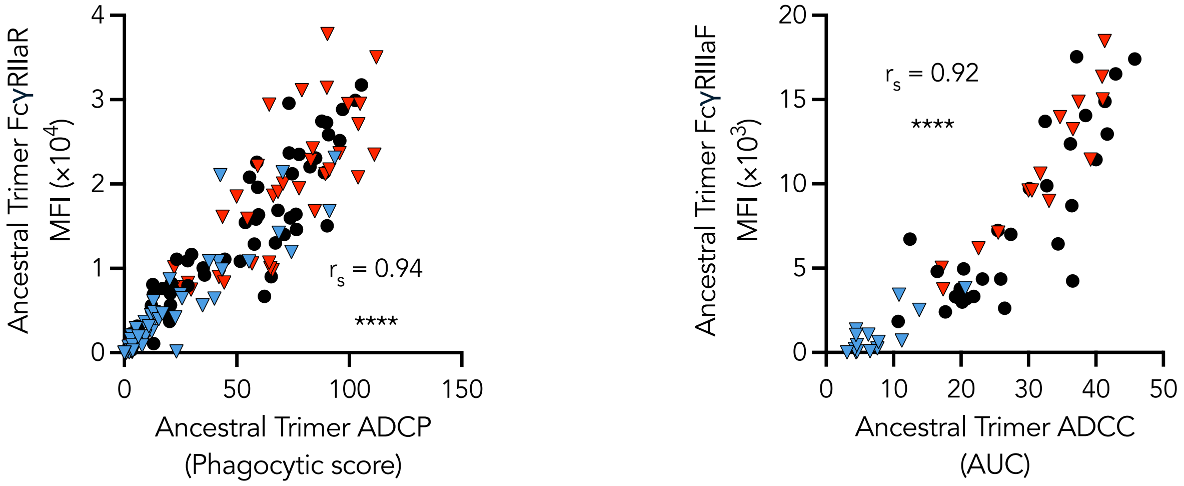


**Supplementary Fig. 3. ADCP and ADCC correlate with engagement of lower-affinity FcγRIIaR and FcγRIIIaF.** Spearman correlation of ADCP with FcγRIIaR binding. Spearman correlation of ADCC with FcγRIIIaF binding. $p < 0.0001$ (****). MFI: median fluorescence intensity. AUC: area under curve.

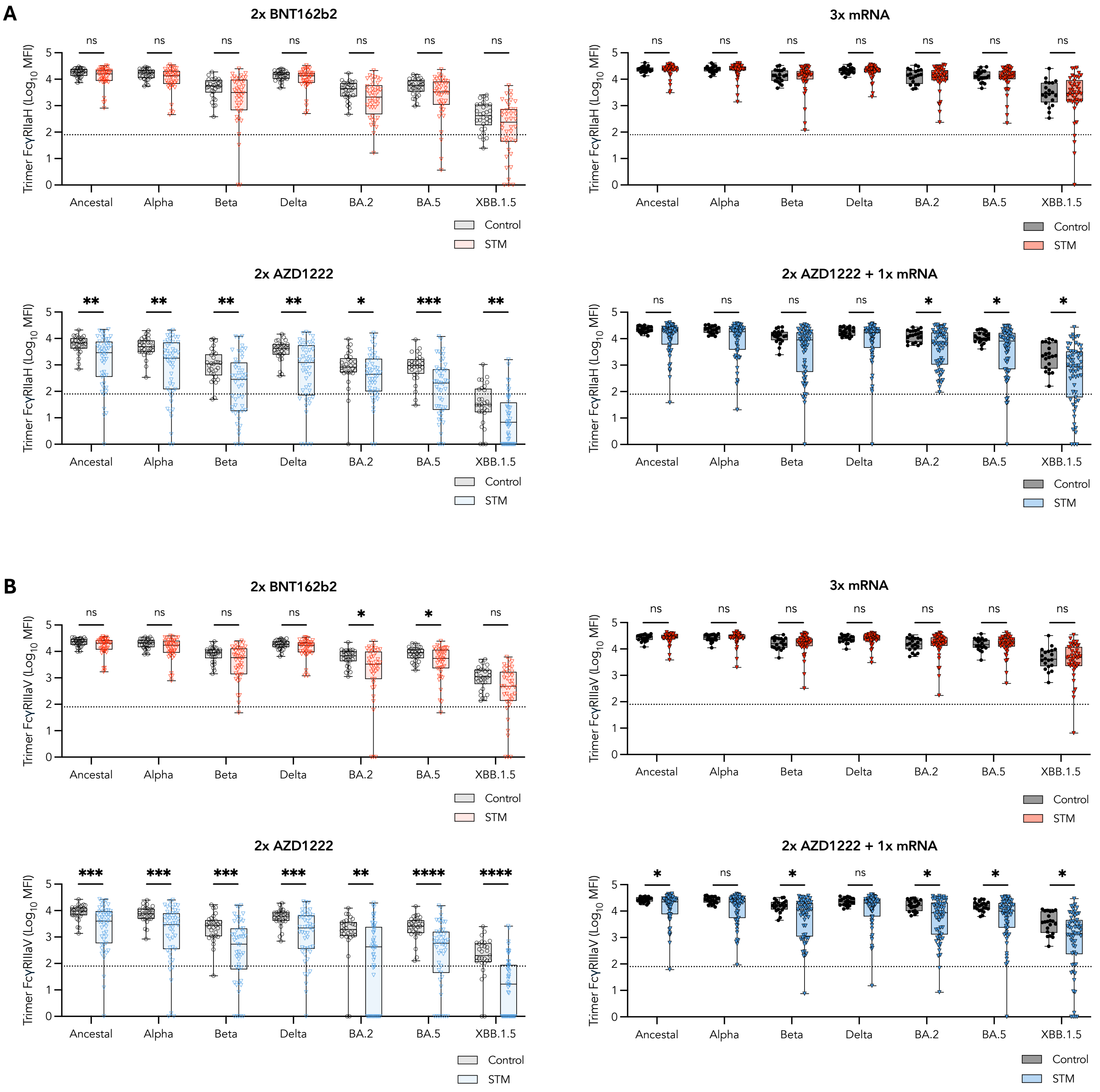


**Supplementary Fig. 4. Robust variant cross-reactive FcγR binding in STM patients.** SARS-CoV-2 ancestral S2-, trimer-, S1-, and RBD- specific engagement of FcγRIIaH and FcγRIIIaV one month post the indicated vaccine regimen in **A)** BNT162b2- or **B)** AZD1222- primed vaccinees. Mann-Whitney *U*-tests performed between STM and control vaccinees within each vaccinee regimen. $p < 0.0001$ (****); $p < 0.001$ (***); $p < 0.01$ (**); $p < 0.05$ (*); non-significant (ns). MFI: median fluorescence intensity.

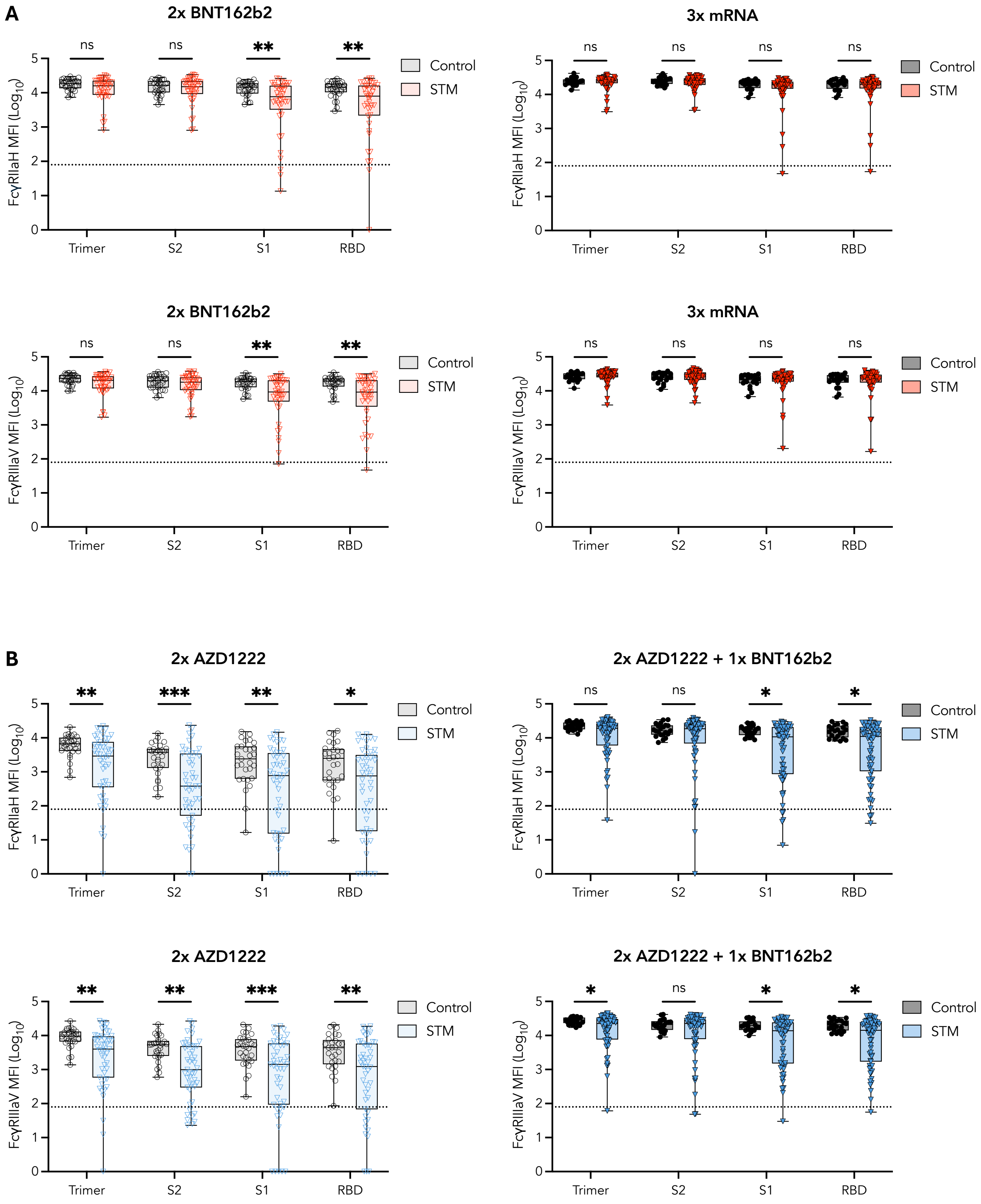


**Supplementary Fig. 5. Robust variant cross-reactive FcγR binding in STM patients.** SARS-CoV-2 variant trimer-specific engagement of **A)** FcγRIIaH and **B)** FcγRIIIaV one month post the indicated vaccine regimen. Mann-Whitney *U*-tests performed between STM and control vaccinees within each vaccinee regimen. $p < 0.0001$ (****); $p < 0.001$ (***); $p < 0.01$ (**); $p < 0.05$ (*); non-significant (ns). MFI: median fluorescence intensity.

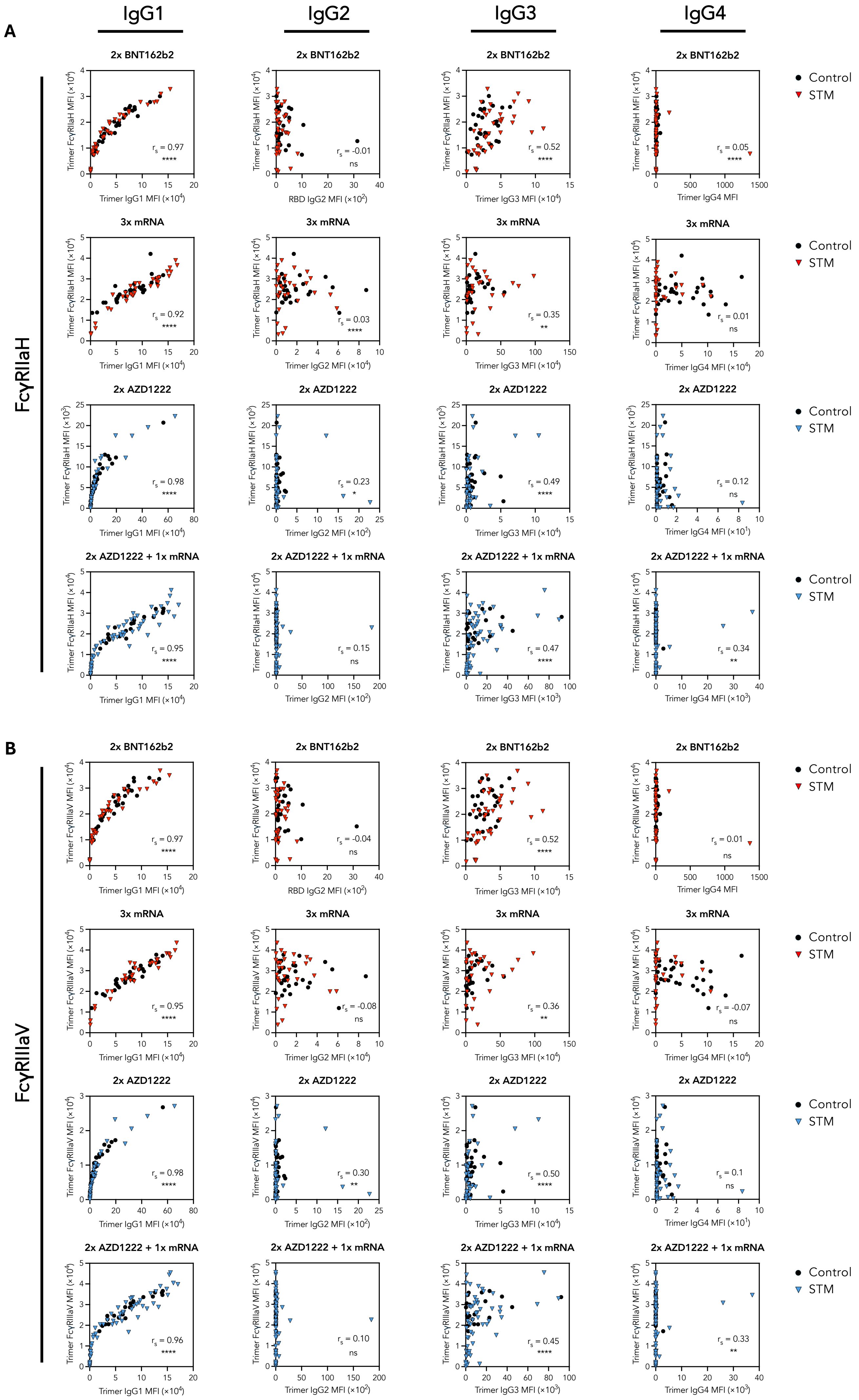

A
IgG1
IgG2
IgG3
IgG4
FcγRIIaH
2x BNT162b2
Trimer FcγRIIaH MFI (×10⁴)
Trimer IgG1 MFI (×10⁴)
RBD IgG2 MFI (×10²)
Trimer IgG3 MFI (×10⁴)
Trimer IgG4 MFI
rs = 0.97 ****
rs = -0.01 ns
rs = 0.52 ****
rs = 0.05 ****
Control
STM
3x mRNA
Trimer IgG2 MFI (×10⁴)
Trimer IgG4 MFI (×10⁴)
rs = 0.92 ****
rs = 0.03 ****
rs = 0.35 **
rs = 0.01 ns
2x AZD1222
Trimer FcγRIIaH MFI (×10³)
Trimer IgG2 MFI (×10²)
Trimer IgG4 MFI (×10¹)
rs = 0.98 ****
rs = 0.23 *
rs = 0.49 ****
rs = 0.12 ns
2x AZD1222 + 1x mRNA
Trimer IgG3 MFI (×10³)
Trimer IgG4 MFI (×10³)
rs = 0.95 ****
rs = 0.15 ns
rs = 0.47 ****
rs = 0.34 **
B
FcγRIIIaV
Trimer FcγRIIIaV MFI (×10⁴)
rs = 0.97 ****
rs = -0.04 ns
rs = 0.52 ****
rs = 0.01 ns
rs = 0.95 ****
rs = -0.08 ns
rs = 0.36 **
rs = -0.07 ns
rs = 0.98 ****
rs = 0.30 **
rs = 0.50 ****
rs = 0.1 ns
rs = 0.96 ****
rs = 0.10 ns
rs = 0.45 ****
rs = 0.33 **

**Supplementary Fig. 6. Trimer-specific IgG1 responses underpin trends in FcγR binding.** Spearman correlations of SARS-CoV-2 trimer-specific **A)** FcγRIIaH and **B)** FcγRIIIaV with IgG1-4 one month post the indicated vaccine regimen. $p < 0.0001$ (****); $p < 0.01$ (**); $p < 0.05$ (*); non-significant (ns). MFI: median fluorescence intensity.

### BNT162b2 prime

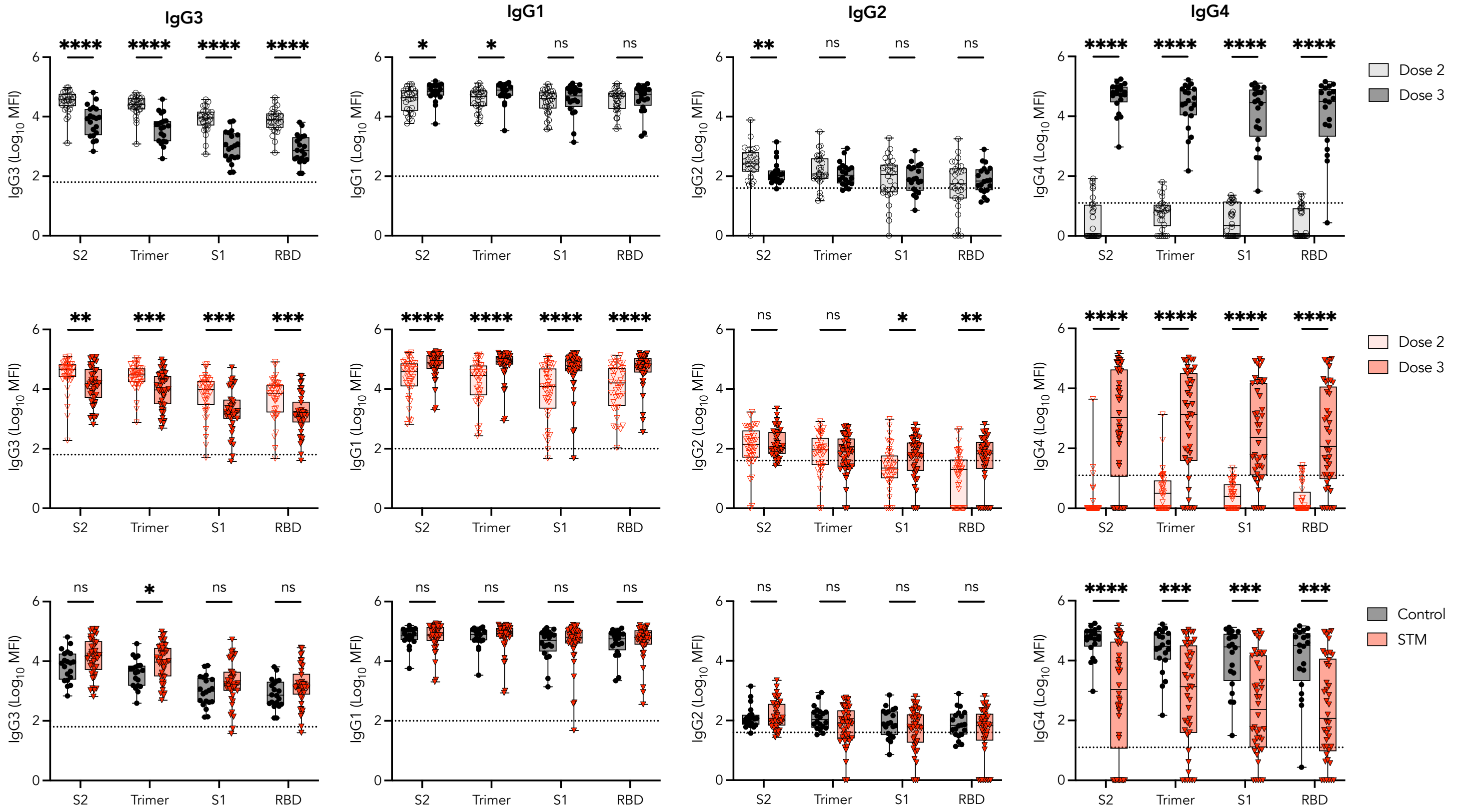


### AZD1222 prime

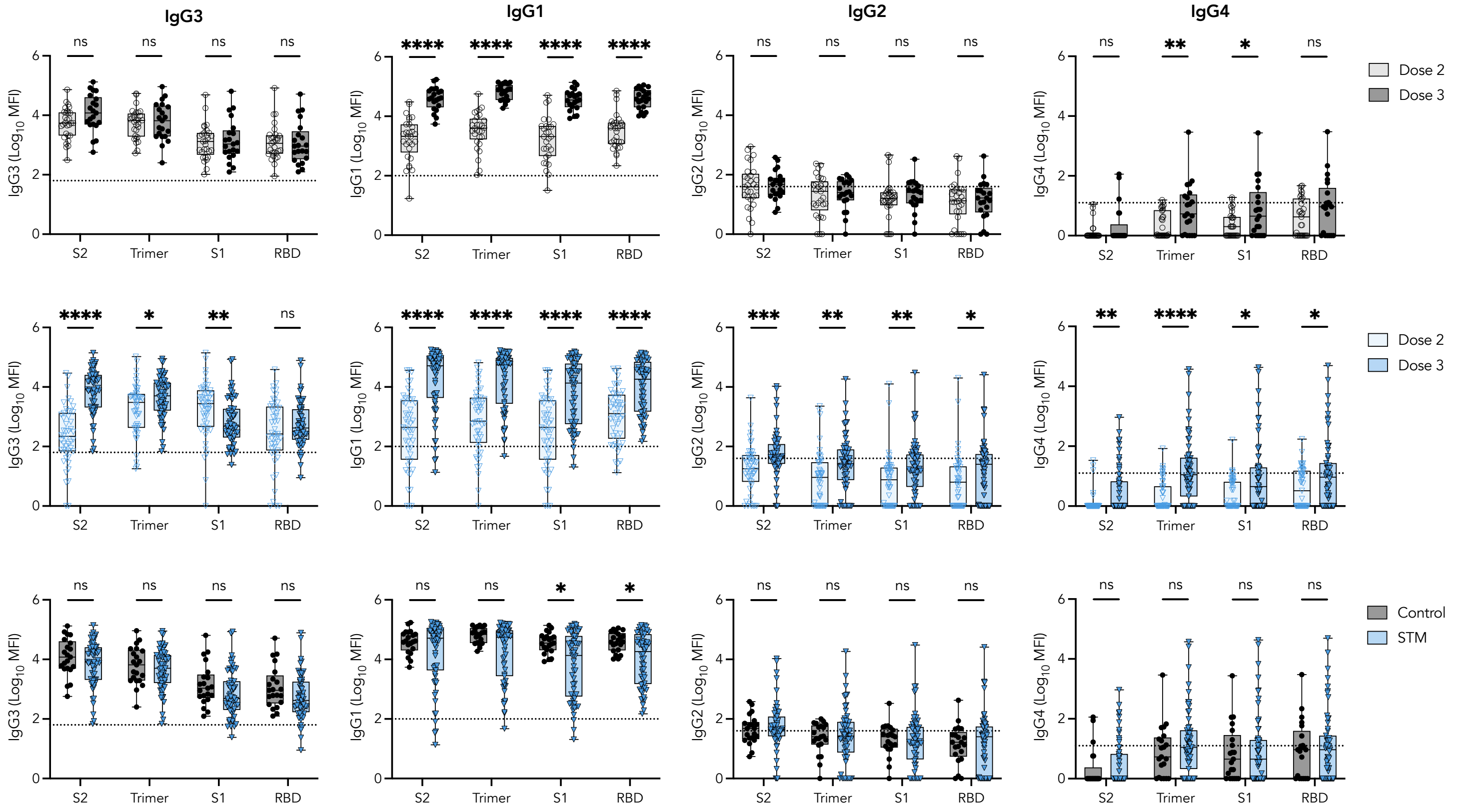

**Supplementary Fig. 7. IgG subclass switching in BNT162b2- and AZD1222- primed patients and controls following a third mRNA vaccination.** SARS-CoV-2 S2-, trimer-, S1-, and RBD-specific IgG1-IgG4 responses one month post dose 2 of the indicated priming regimen and one month post mRNA-based third vaccination. Mann-Whitney *U*-tests performed between STM and control vaccinees or between dose 2 and dose 3 responses within each vaccinee regimen. $p < 0.0001$ (****); $p < 0.001$ (***); $p < 0.01$ (**); $p < 0.05$ (*); non-significant (ns). MFI: median fluorescence intensity.

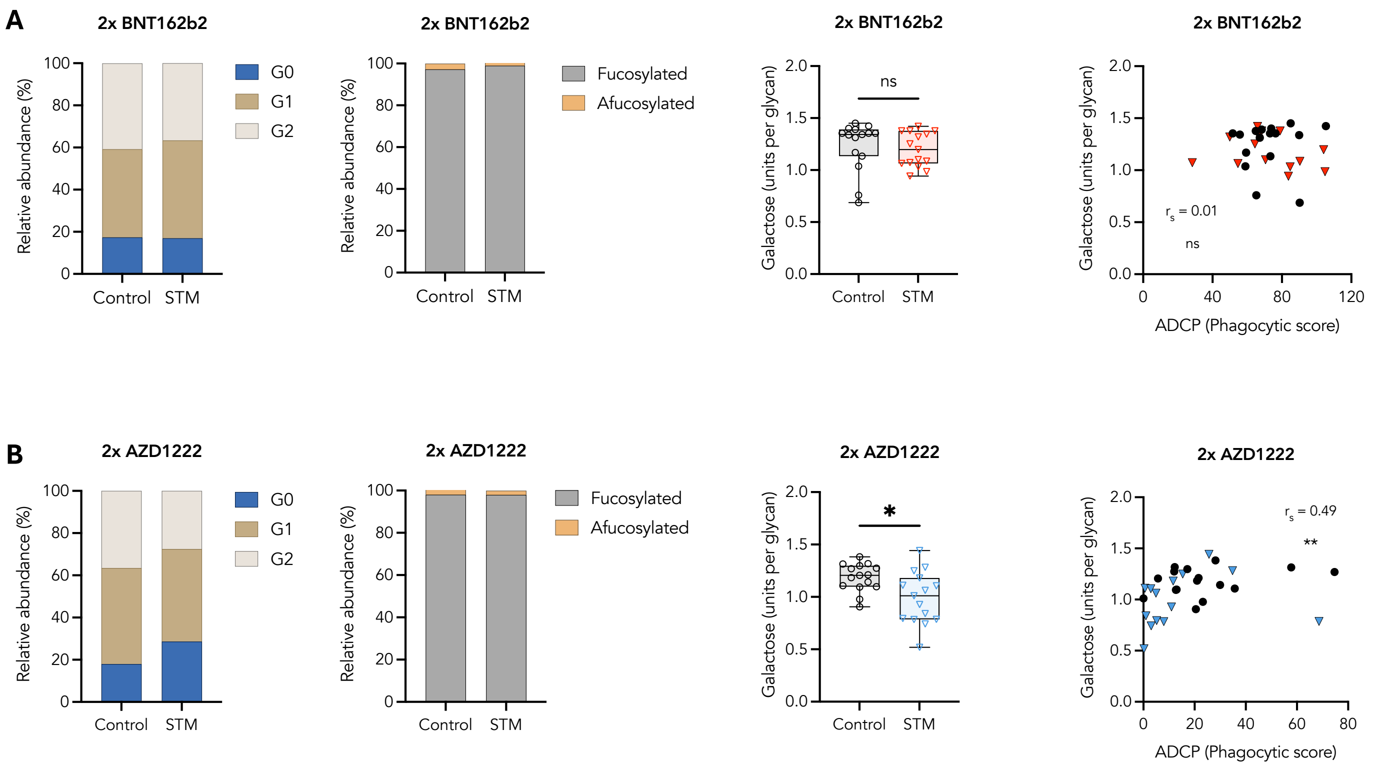


**Supplementary Fig. 8. Modulation of antigen-specific IgG glycosylation profiles and association with Fc effector function following COVID-19 vaccination.** RBD-specific IgG galactosylation and fucosylation and Spearman correlations between galactosylation and ADCP one month post two **A)** BNT162b2 doses ($n$ = 15 control; $n$ = 15 STM) or **B)** AZD1222 doses ($n$ = 15 control; $n$ = 15 STM). Mann-Whitney $U$-tests performed between STM and control vaccinees within each vaccinee regimen. $p < 0.01$ (**); $p < 0.05$ (*); non-significant (ns).

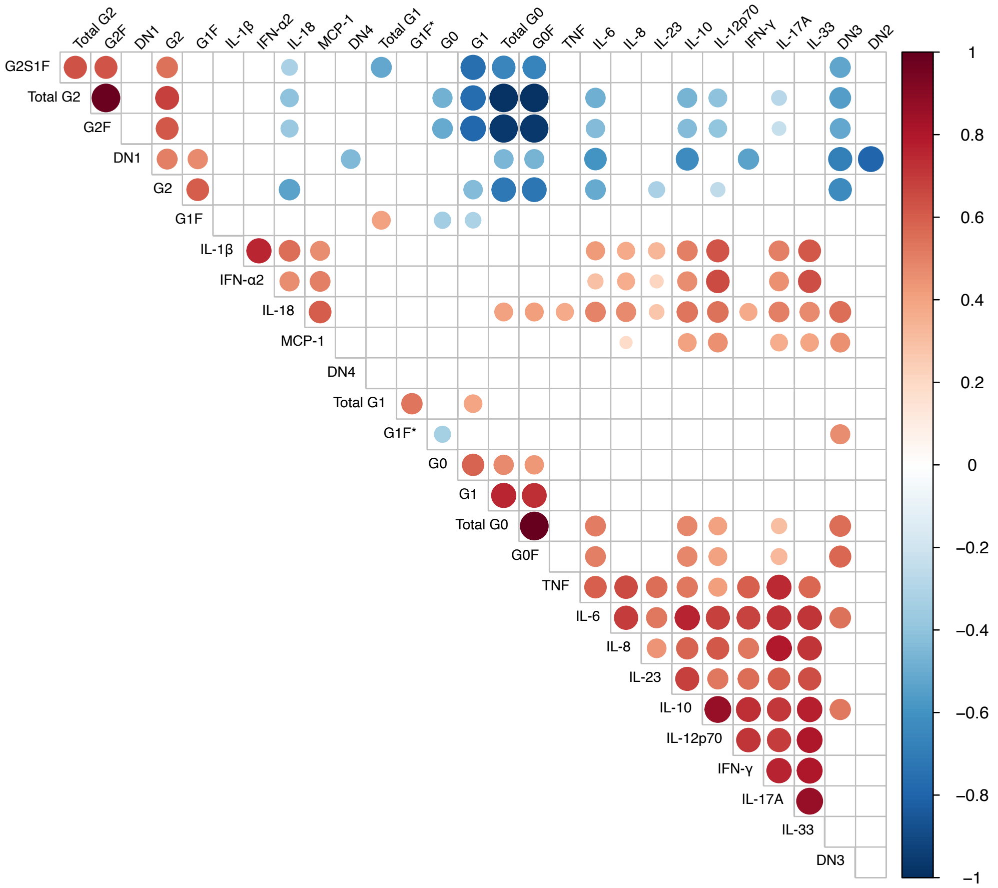


**Supplementary Fig. 9. Correlations between inflammatory biomarkers.** Correlation matrix showing Spearman correlations between inflammatory biomarkers in control and STM vaccinees one month post two BNT162b2 doses. Statistical significance was determined with an unpaired, two-sided *t*-test with Benjamini–Hochberg adjustment. Size of each circle indicates magnitude of the correlation coefficient. Only significant correlations are shown; non-significant correlations are indicated as white boxes.

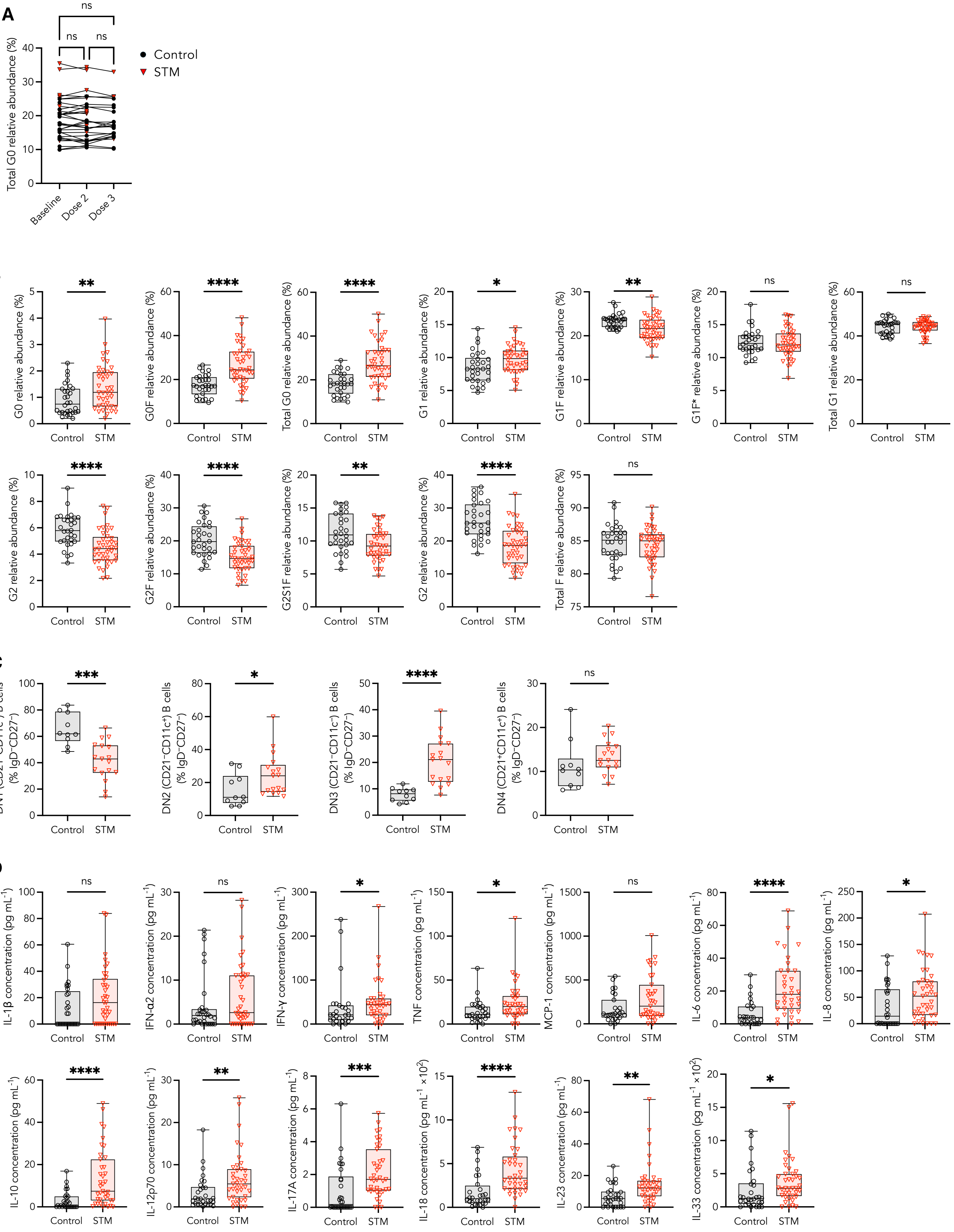
A
B
C
D
Control
STM
Total G0 relative abundance (%)
Baseline
Dose 2
Dose 3
ns
G0 relative abundance (%)
G0F relative abundance (%)
Total G0 relative abundance (%)
G1 relative abundance (%)
G1F relative abundance (%)
G1F* relative abundance (%)
Total G1 relative abundance (%)
G2 relative abundance (%)
G2F relative abundance (%)
G2S1F relative abundance (%)
G2 relative abundance (%)
Total F relative abundance (%)
DN1 (CD21+CD11c−) B cells (% IgD−CD27−)
DN2 (CD21−CD11c+) B cells (% IgD−CD27−)
DN3 (CD21−CD11c−) B cells (% IgD−CD27−)
DN4 (CD21+CD11c+) B cells (% IgD−CD27−)
IL-1β concentration (pg mL−1)
IFN-α2 concentration (pg mL−1)
IFN-γ concentration (pg mL−1)
TNF concentration (pg mL−1)
MCP-1 concentration (pg mL−1)
IL-6 concentration (pg mL−1)
IL-8 concentration (pg mL−1)
IL-10 concentration (pg mL−1)
IL-12p70 concentration (pg mL−1)
IL-17A concentration (pg mL−1)
IL-18 concentration (pg mL−1 ×10²)
IL-23 concentration (pg mL−1)
IL-33 concentration (pg mL−1 ×10²)

**Supplementary Fig. 10. Inflammatory biomarkers in STM patients and control vaccinees.** **A)** Abundance of total agalactosylated (G0) IgG glycoforms over the course of a three-dose BNT162b2 vaccination regimen. Baseline: pre-vaccination; Dose 2: one month post second dose; Dose 3: one month post third dose. **B)** Relative abundance of IgG glycoforms, **C)** relative frequencies of DN memory B cell subsets, and **D)** concentration of 13 cytokines one month post second BNT162b2 vaccination. Kruskal-Wallis test performed to assess changes in IgG glycosylation across timepoints. Mann-Whitney *U*-tests performed between STM and control vaccinees. $p < 0.0001$ (****); $p < 0.001$ (***); $p < 0.01$ (**); $p < 0.05$ (*); non-significant (ns). MFI: median fluorescence intensity.

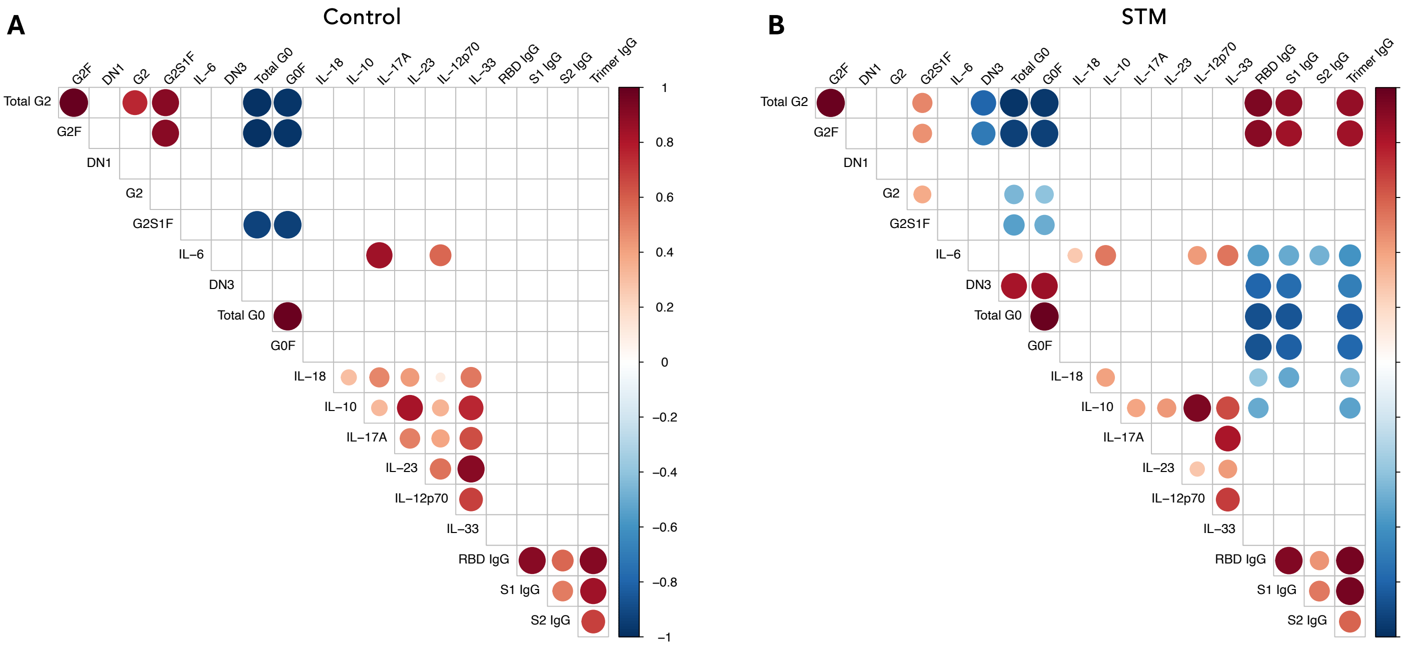


**Supplementary Fig. 11. Correlations between inflammatory biomarkers and SARS-CoV-2-specific IgG. A-B)** Correlation matrices showing the Spearman correlations between SARS-CoV-2-specific IgG responses and differentially abundant inflammatory biomarkers for **A)** control and **B)** STM vaccinees one month post two BNT162b2 doses. Statistical significance was determined with an unpaired, two-sided *t*-test with Benjamini–Hochberg adjustment. Size of each circle indicates magnitude of the correlation coefficient. Only significant correlations are shown; non-significant correlations are indicated as white boxes.

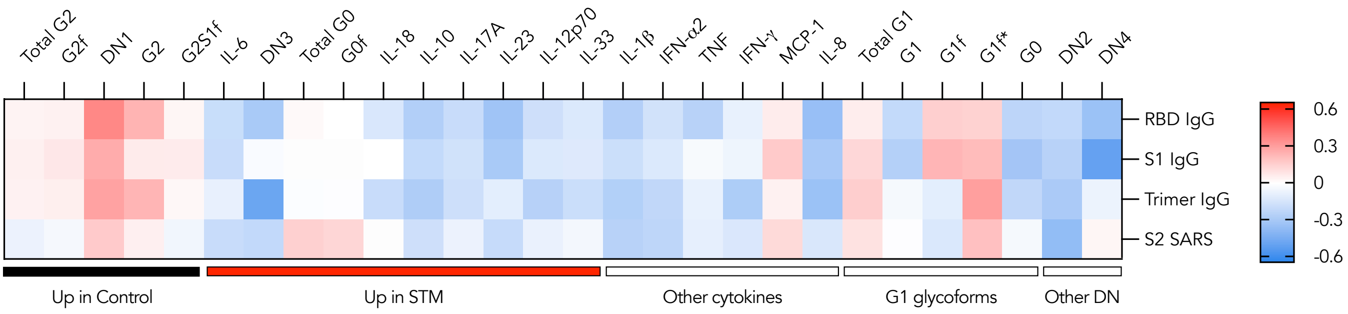


**Supplementary Fig. 12. Correlations between inflammatory biomarkers and SARS-CoV-2-specific IgG one month post third mRNA vaccination.** Spearman correlations between differentially abundant inflammatory biomarkers and SARS-CoV-2-specific IgG responses for control and STM vaccinees one month post third mRNA vaccination. Statistical significance adjusted for multiple comparisons via Holm-Šídák method. No significant correlations were identified.
